# Risk factors for long-term consequences of COVID-19 in hospitalised adults in Moscow using the ISARIC Global follow-up protocol: StopCOVID cohort study

**DOI:** 10.1101/2021.02.17.21251895

**Authors:** Daniel Munblit, Polina Bobkova, Ekaterina Spiridonova, Anastasia Shikhaleva, Aysylu Gamirova, Oleg Blyuss, Nikita Nekliudov, Polina Bugaeva, Margarita Andreeva, Audrey DunnGalvin, Pasquale Comberiati, Christian Apfelbacher, Jon Genuneit, Sergey Avdeev, Valentina Kapustina, Alla Guekht, Victor Fomin, Andrey A Svistunov, Peter Timashev, Thomas M Drake, Sarah Wulf Hanson, Laura Merson, Peter Horby, Louise Sigfrid, Janet T Scott, Malcolm G Semple, John O Warner, Theo Vos, Piero Olliaro, Petr Glybochko, Denis Butnaru, Sechenov StopCOVID Research Team

## Abstract

**Background:** The long-term sequalae of COVID-19 remain poorly characterised. In this study, we aimed to assess long-standing symptoms (LS) (symptoms lasting from the time of discharge) in previously hospitalised patients with COVID-19 and assess associated risk factors.

**Methods:** This is a longitudinal cohort study of adults (≥18 years of age) with clinically diagnosed or laboratory-confirmed COVID-19 admitted to Sechenov University Hospital Network in Moscow, Russia. Data were collected from patients discharged between April 8 and July 10, 2020. Participants were interviewed via telephone using Tier 1 ISARIC Long-term Follow-up Study CRF and the WHO CRF for Post COVID conditions. Reported symptoms were further categorised based on the system(s) involved. Additional information on dyspnoea, quality of life and fatigue was collected using validated instruments. Multivariable logistic regressions were performed to investigate risk factors for development of LS categories.

**Findings:** Overall, 2,649 of 4,755 patients discharged from the hospitals were available for the follow-up and included in the study. The median age of the patients was 56 years (IQR, 46–66) and 1,353 (51.1%) were women. The median follow-up time since hospital discharge was 217.5 (200.4-235.5) days. At the time of the follow-up interview 1247 (47.1%) participants reported LS. Fatigue (21.2%, 551/2599), shortness of breath (14.5%, 378/2614) and forgetfulness (9.1%, 237/2597) were the most common LS reported. Chronic fatigue (25%, 658/2593) and respiratory (17.2% 451/2616) were the most common LS categories. with reporting of multi-system involvement (MSI) less common (11.3%; 299). Female sex was associated with LS categories of chronic fatigue with an odds ratio of 1.67 (95% confidence interval 1.39 to 2.02), neurological (2.03, 1.60 to 2.58), mood and behaviour (1.83, 1.41 to 2.40), dermatological (3.26, 2.36 to 4.57), gastrointestinal (2.50, 1.64 to 3.89), sensory (1.73, 2.06 to 2.89) and respiratory (1.31, 1.06 to 1.62). Pre-existing asthma was associated with neurological (1.95, 1.25 to 2.98) and mood and behavioural changes (2.02, 1.24 to 3.18) and chronic pulmonary disease was associated with chronic fatigue (1.68, 1.21 to 2.32).

**Interpretation:** 6 to 8 months after acute infection episode almost a half of patients experience symptoms lasting since hospital discharge. One in ten individuals experiences MSI. Female sex is the main risk factor for majority of the LS categories. chronic pulmonary disease is associated with a higher risk of chronic fatigue development, and asthma with neurological and mood and behaviour changes. Individuals with LS and MSI should be the main target for future research and intervention strategies.

**Funding:** This study is supported by Russian Fund for Basic Research and UK Embassy in Moscow. The ISARIC work is supported by grants from: the NIHR Health Protection Research Unit (HPRU) in Emerging and Zoonotic Infections at University of Liverpool in partnership with Public Health England (PHE), in collaboration with Liverpool School of Tropical Medicine and the University of Oxford [award 200907], Wellcome Trust and Department for International Development [215091/Z/18/Z], and the Bill and Melinda Gates Foundation [OPP1209135], EU Platform for European Preparedness Against (Re-) emerging Epidemics (PREPARE) [FP7 project 602525] This research was funded in part, by the Wellcome Trust. The views expressed are those of the authors and not necessarily those of the DID, NIHR, Wellcome Trust or PHE.

**Research in context:** *Evidence before this study:* Evidence suggests that COVID-19 may result in short- and long-term consequences to health. Most studies do not provide definitive answers due to a combination of short follow-up (2-3 months), small sample size, and use of non-standardised tools. There is a need to study the longer-term health consequences of previously hospitalised patients with COVID-19 infection and to identify risk factors for sequalae.

*Added value of this study:* To our knowledge, this is the largest cohort study (n=2,649) with the longest follow-up since hospital discharge (6-8 months) of previously hospitalised adult patients. We found that 6-8 months after discharge from the hospital, around a half (47.1%) of patients reported at least one long-standing symptom since discharge. Once categories of symptoms were assessed, chronic fatigue and respiratory problems were the most frequent clusters of long-standing symptoms in our patients. Of those patients having long-term symptoms, a smaller proportion (11.3%) had multisystem involvement, with three or more categories of long-standing symptoms present. Although most patients developed symptoms since discharge, a smaller number of individuals experienced symptom beginning symptom appearing weeks or months after the acute phase. Female sex was a predictor for most of the symptom categories at the time of the follow-up interview, with chronic pulmonary disease associated with chronic fatigue-related symptoms, and asthma with a higher risk of neurological symptoms, mood and behaviour problems.

*Implications of all the available evidence:* The majority of patients experienced long-lasting symptoms 6 to 8 months after hospital discharge and almost half reported at least one long-standing symptom, with chronic fatigue and respiratory problems being the most frequent. A smaller number reported multisystem impacts with three or more long-standing categories present at follow-up. A higher risk was found for women, for chronic pulmonary disease with chronic fatigue, and neurological symptoms and mood and behaviour problems with asthma. Patterns of the symptom development following COVID-19 should be further investigated in future research.

## INTRODUCTION

The emergence of severe acute respiratory syndrome coronavirus 2 (SARS-CoV-2) has placed a significant burden on health services and society world-wide. There have now been well over 100 million coronavirus disease 2019 (COVID-19) cases reported with a mortality rate of around 2.2%.”globally ^1^. The acute presentation of COVID-19 has now been well investigated, with fever, cough, shortness of breath and anosmia among the most commonly reported symptoms ^2-4^.

It has become evident that a substantial proportion of people experience ongoing symptoms including fatigue and muscle weakness, joint and muscle pain, and breathlessness, months after the acute phase of COVID-19 ^5-7^. This phenomenon is now commonly referred to as Long COVID but has also been described as post-COVID syndrome, the post-COVID-19 condition^8^ or patients have been labelled COVID long-haulers ^9,10^. The recently published UK National Institute for Health and Clinical Excellence (NICE) COVID-19 guideline defines the syndrome as “signs and symptoms that develop during or after an infection consistent with COVID-19, continue for more than 12 weeks and are not explained by an alternative diagnosis” ^11^. However, publications have used a range of definitions based on limited evidence from small, heterogeneous cohorts ^7^. Overall, there is still a paucity of long-term follow-up data, which means we have limited knowledge of the full range of symptoms, duration of disease and potential risk factors. Recently published data from China describing long-term consequences of COVID-19 show that 76% of previously hospitalised adult patients have at least one symptom 6 months after acute infection ^6^.

There is an urgent need for accurate long-term follow-up of COVID-19 patients ^7^, to inform future management plans and address the devastating impacts of this condition on the quality of life (QoL) of people affected. This observational cohort study aimed to investigate the incidence of long-term consequences in adults previously hospitalised for COVID-19 and to assess risk factors for Long COVID in Moscow, Russia. We used the standardised follow-up data collection protocol of the International Severe Acute Respiratory and Emerging Infection Consortium (ISARIC).

## METHODS

### Study design, setting and participants

This is a longitudinal cohort study of patients with suspected or confirmed COVID-19 infection admitted to four Sechenov University Hospital Network in Moscow, Russia. We collected the follow-up data between December 2, 2020 and January 14, 2021 from patients discharged between April 8, 2020 and July 10, 2020. We included adult patients (≥18 years of age), with both reverse transcriptase polymerase chain reaction (RT-PCR) confirmed SARS-CoV-2 infection, corresponding to U07.1 as per International Classification of Diseases (ICD), and clinically confirmed infection, encoded as U07.2 when the laboratory testing result is negative, inconclusive or unavailable.

The acute phase data were extracted from electronic medical records (EMR) and the Local Health Information System (HIS) at the host institution using the modified and translated ISARIC WHO Clinical Characterisation protocol (CCP) ^12^. Details of the acute phase data collection are described elsewhere ^3^. The acute-phase dataset included demographics, symptoms, comorbidities, chest computer tomography (CT) reports, supportive care data, and clinical outcomes at discharge. The study was approved by the Sechenov University Local Ethics Committee on April 22, 2020 (protocol number 08–20). A protocol amendment enabling serial follow-up of the cohort was approved on November 13, 2020.

Information about the current condition and persisting symptoms was collected by telephone using the Tier 1 ISARIC Long-term Follow-up Study case report form (CRF) developed by the ISARIC Global COVID-19 follow up working group, with adaptations, translated into Russian assessing the patients’ physical and mental health ^9^. Additional information was added from the WHO CRF for Post COVID conditions ^13^. The follow-up CRF documented data on demographics, lifestyle and socioeconomic data, history of vaccination, hospital stay and readmissions, mortality (after the initial index event), presence of newly developed symptoms at the time of follow-up interview and symptom duration (1-2 weeks; > 2-4 weeks; >1-2 months; >2-3 months; >3-6 months; from the time of discharge), dyspnoea, fatigue, quality of life (QoL) and difficulties in functioning before the COVID-19 illness and at the time of the follow-up (**Supplementary material**).

The participants were asked to report on dyspnoea, QoL and difficulties in functioning before the COVID-19 illness (retrospectively) and presently (at the time they were completing the questionnaire), to enable comparative assessment of the respective variables. We used the British Medical Research Council (MRC) dyspnoea scale, the EuroQoL five-dimension five-level (EQ-5D-5L) questionnaire, the EuroQoL Visual Analogue Scale (EQ-VAS) asking participants to score their QoL from 0 (worst imaginable health) to 100 (best imaginable health), UNICEF/Washington disability score and World Health Organisation Disability Assessment Schedule (WHODAS 2.0). The study was registered with EuroQoL as part of the ISARIC collaborative effort (EuroQoL ID 37035).

Data collection and entry were performed by a team of medical students who signed confidentiality agreements, underwent training in basic data entry into REDCap and telephone interviews. Students have already had extensive data extraction experience gained from the previous research ^3^ and were supervised by senior academic staff members.

The research team members attempted to contact patients three times before declaring them lost to follow up. If available by telephone, the patients were asked to provide their verbal consent to the interview. The patient’s record ID was cross-checked with existing demographic data to ensure reliable collation of the follow-up surveys with the existing acute-phase data extracted previously.

### Data management

We used REDCap electronic data capture tools (Vanderbilt University, Nashville, TN, USA) hosted at Sechenov University and Microsoft Excel (Microsoft Corp, Redmond, WA, USA) for data collection, storage and management ^11,12^. The baseline characteristics, including demographics, symptoms on admission and comorbidities had been extracted from EMRs and entered into REDCap previously.

### Definitions

The acute disease severity was stratified in accordance with Arnold et al. ^10^ by a three-category scale based on the degree of required supportive care during hospital stay: mild (no supplementary oxygen or intensive care), moderate (supplementary oxygen during hospitalisation) and severe (need for non-invasive respiratory modalities (NIV), invasive mechanical ventilation (IMV) and/or admission to intensive care unit (ICU)). A difference of 10 points at EQ-VAS defined relevant change in the health status ^4^.

For the purpose of this study, we defined “long-standing symptoms” (LS) as symptoms present since hospital discharge. LS present at the time of follow-up were also categorised into respiratory, gastrointestinal, dermatological, chronic fatigue, neurological, mood and behaviour, sensory (**Table S1**). Symptom categorisation was based on previously published literature ^14,15^ and international expert group discussions.

### Outcomes

The primary outcomes included the prevalence of LS and prevalence of categories of LS at follow-up.

The secondary outcomes included any symptom prevalence at follow-up (regardless of their duration), and health status assessed by EuroQoL VAS, fatigue scale, and MRC dyspnea scale.

### Statistical analysis

Descriptive statistics were calculated for baseline characteristics. Continuous variables were summarised as median (with interquartile range) and categorical variables as frequency (percentage). The chi-squared test or Fisher’s exact test was used for testing differences in proportions between groups. The Wilcoxon rank-sum test was used for testing the hypotheses about differences in means between the groups.

We performed multivariable logistic regression to investigate associations of demographic characteristics, comorbidities, and severity of acute phase COVID-19 with LS categories presence at the time of the follow-up interview. To enhance the robustness of the effect estimates, only comorbidities that were present in at least 3% of the cohort were included in the modelling. Primary analysis was performed using the full dataset, whereas sensitivity analysis included only a subset of people with RT-PCR confirmed SARS-CoV-2 infection (ICD U07.1). We have previously found no significant differences in clinical signs, symptoms, laboratory test results and risk factors for in-hospital mortality between clinically diagnosed patients and patients with positive RT-PCR ^3^. Therefore, primary analysis was performed using the full cohort. Robustness of findings was then investigated via sensitivity analysis which included only a subset with confirmed SARS-CoV-2 infection. We have not performed any imputation for missing data.

Venn diagrams were used to present the coexistence of the five most common long-standing symptoms.

Two-sided p-values were reported for all statistical tests, a p-value below 0.05 was considered to be statistically significant. Statistical analysis was performed using R version 3.5.1.

## RESULTS

As outlined in **Figure 1**, out of 5,040 patients hospitalised with suspected COVID-19 to the hospitals before July 10, 2020, 4,755 were discharged alive or transferred to another facility. Out of 4,019 patients with accurate contact information available, 2,649 were available for follow-up (response rate 68.5%), 2,649 of whom had no missing baseline data in the electronic database and were included in the analysis. Of the 3,868 patients with contact information available 52 (1.3%) died after the hospital discharge.

**Figure 1.**
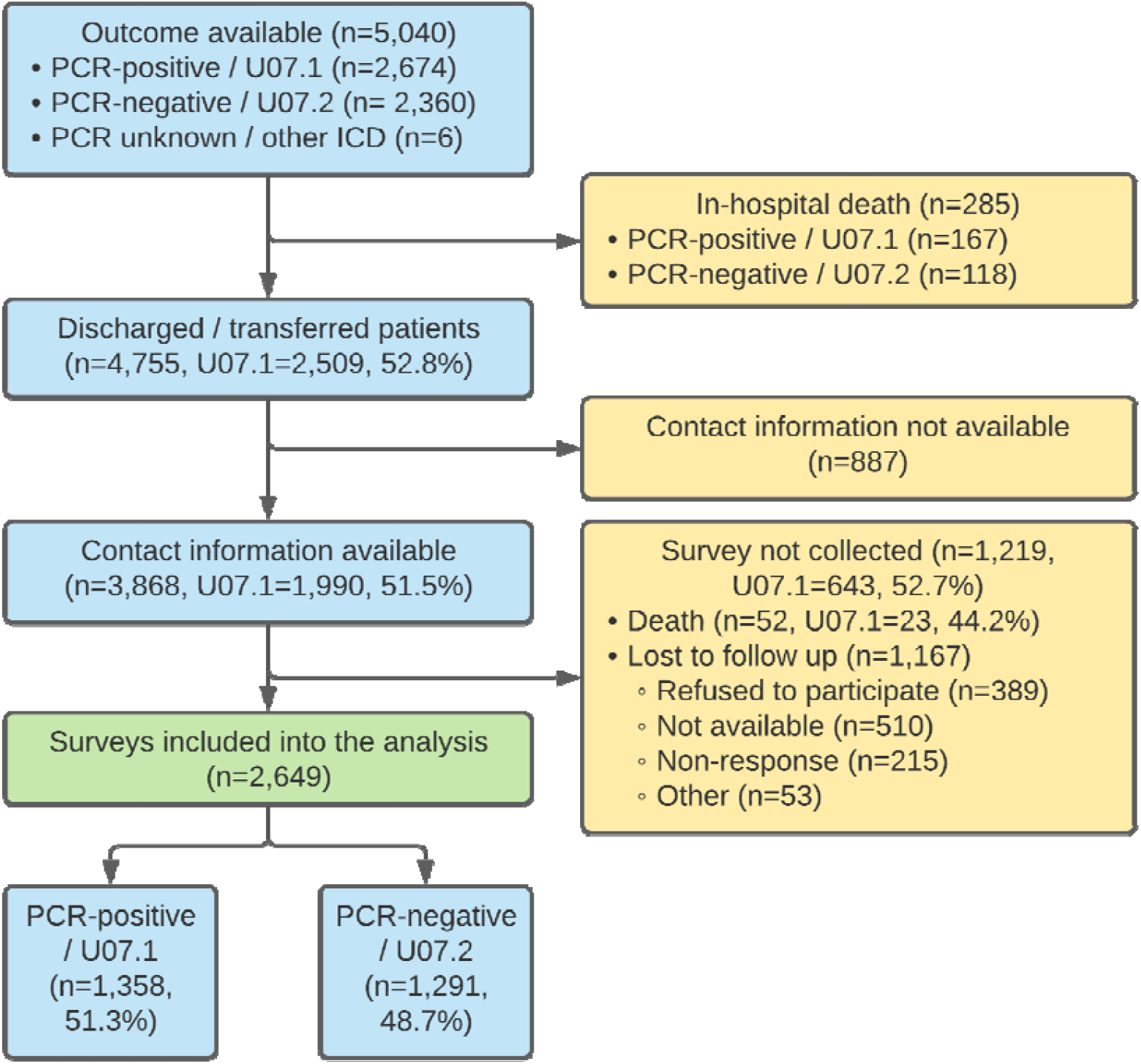
Flow diagram of patients with COVID-19 admitted to Sechenov University Hospital Network between April 8 and July 10, 2020. PCR, polymerase chain reaction.

Out of 2,649 participants, 1,358 patients (51.3%) had RT-PCR-confirmed SARS-CoV-2 infection, whereas 1291 (48.7%) were clinically diagnosed with COVID-19. In-hospital case fatality ratio was 167/2674 (6.2%) in laboratory-confirmed and 118/2360 (5%) in clinically diagnosed patients (p=0.09). The median age was 56 years (IQR, 46–66; range, 18–100 years), 1,353 (51.1%) were women. Median follow-up time post-discharge was 217.5 days (IQR 200.4-235.5, range 18-100). 1,948 participants (77.4%) had a higher education; 1,531, (59.8%) of the participants were working part- or full-time and 830 (32.4%) were retired (**Table 1**).

**Table 1.** Demographic characteristics of patients admitted to the Sechenov University Hospital Network. Data are n (%), n/N (%), or median (IQR). Statistically significant results (p<0.05) are highlighted in bold.

|  |  | Total | Laboratory-confirmed<br>RT-PCR “+” (n=1358) | Clinically diagnosed<br>RT-PCR “-” (n=1291) |
| --- | --- | --- | --- | --- |
| Age (median, IQR) |  | 56 (46-66) | 57 (47-67) | 55 (44-65) |
| Age groups (years) | <50 | 870 (32.8%) | 407 (30%) | 463 (35.9%) |
|  | 50–69 | 1294 (48.8%) | 673 (49.6%) | 621 (48.1%) |
|  | 70–79 | 304 (11.5%) | 169 (12.4%) | 135 (10.5%) |
|  | ≥80 | 181 (6.8%) | 109 (8%) | 72 (5.6%) |
| Sex | Female | 1353 (51.1%) | 683 (50.3%) | 670 (51.9%) |
| Time from admission to discharge, days | Median (IQR) | 14.6 (12-17.6) | 15.1 (12.8-18.7) | 13.7 (11.6-16.1) |
| Time since discharge, days | Median (IQR) | 217.5<br>(200.4-235.5) | 215.3<br>(195.5-234.5) | 220.5<br>(204.4-237.3) |
| Highest completed educational level | School | 240 (9.5%) | 125 (9.7%) | 115 (9.3%) |
|  | University | 1948 (77.4%) | 989 (76.8%) | 959 (78%) |
|  | Not completed formal education or training | 37 (1.5%) | 13 (1%) | 24 (2%) |
| **Occupation/ Working status** |  |  |  |  |
| Occupation/working status before Covid-19 | Working full-time | 1473 (57.5%) | 745 (54.9%) | 728 (56.4%) |
|  | Working part-time | 58 (2.3%) | 25 (1.8%) | 33 (2.6%) |
|  | Full time carer (children or others) | 47 (1.8%) | 22 (1.6%) | 25 (1.9%) |
|  | Unemployed | 47 (1.8%) | 23 (1.7%) | 24 (1.9%) |
|  | Unable to work due to chronic illness | 5 (0.2%) | 2 (0.1%) | 3 (0.2%) |
|  | Student | 21 (0.8%) | 8 (0.6%) | 13 (1%) |
|  | Retired/ Early retirement due to illness | 871 (34.0%) | 459 (33.8%) | 412 (31.9%) |
|  | Do not want to answer | 155 (6.1%) | 82 (6%) | 73 (5.7%) |
| **Comorbidities** |  |  |  |  |
| Chronic cardiac disease (not hypertension) |  | 476 (18.1%) | 273 (20.3%) | 203 (15.8%) |
| Hypertension |  | 1219 (46.2%) | 646 (47.8%) | 573 (44.4%) |
| Revascularization of peripheral and/or coronary arteries |  | 98 (3.7%) | 68 (5.1%) | 30 (2.3%) |
| Chronic pulmonary disease (not asthma) |  | 193 (7.3%) | 110 (8.2%) | 83 (6.5%) |
| Asthma (physician diagnosed) |  | 121 (4.6%) | 73 (5.4%) | 48 (3.7%) |
| Chronic kidney disease |  | 123 (4.7%) | 72 (5.4%) | 51 (4%) |
| Obesity (as defined by clinical staff) |  | 514 (19.6%) | 263 (19.6%) | 251 (19.6%) |
| Moderate or severe liver disease |  | 19 (0.7%) | 11 (0.8%) | 8 (0.6%) |
| Mild liver disease |  | 50 (1.9%) | 24 (1.8%) | 26 (2%) |
| Asplenia |  | 8 (0.3%) | 4 (0.3%) | 4 (0.3%) |
| Chronic neurological disorder |  | 121 (4.6%) | 78 (5.8%) | 43 (3.4%) |
| Malignant neoplasm |  | 93 (3.5%) | 62 (4.6%) | 31 (2.4%) |
| Chronic hematologic disease |  | 30 (1.1%) | 21 (1.6%) | 9 (0.7%) |
| AIDS / HIV | on ART | 4 (0.2%) | 0 (0%) | 4 (0.3%) |
|  | not on ART | 5 (0.2%) | 3 (0.2%) | 2 (0.2%) |
| Diabetes Mellitus | Type 1 | 13 (0.5%) | 6 (0.4%) | 7 (0.5%) |
|  | Type 2 | 369 (14.1%) | 198 (14.7%) | 171 (13.4%) |
| Rheumatologic disorder |  | 96 (3.7%) | 43 (3.2%) | 53 (4.1%) |
| Dementia |  | 25 (1%) | 18 (1.3%) | 7 (0.5%) |
| Tuberculosis |  | 2 (0.1%) | 1 (0.1%) | 1 (0.1%) |
| Malnutrition |  | 6 (0.2%) | 2 (0.1%) | 4 (0.3%) |
| With no comorbidities |  | 908 (34.3%) | 452 (33.3%) | 456 (35.3%) |
| With 1 comorbidity |  | 723 (27.3%) | 341 (25.1%) | 382 (29.6%) |
| With 2 comorbidities |  | 512 (19.3%) | 267 (19.7%) | 245 (19%) |
| With 3+ comorbidities |  | 506 (19.1%) | 298 (21.9) | 208 (16.1%) |
| **COVID-19 Severity** |  |  |  |  |
|  | Mild (not requiring supplementary oxygen or ventilation) | 1679 (63.4%) | 841 (61.9%) | 838 (64.9%) |
|  | Moderate (requiring only supplementary oxygen) | 902 (34%) | 479 (35.3%) | 423 (32.8%) |
|  | Severe (requiring non-invasive ventilation, or invasive ventilation or intensive care admission) | 68 (2.6%) | 38 (2.8%) | 30 (2.3%) |

The most common pre-existing comorbidity on admission was hypertension (1,219, 46.2%), followed by obesity (514, 19.6%) and type II diabetes (369, 14.1%). Most of the patients had mild COVID-19 (1,637, 63.2%), with 902 (34%) classified as moderate and 68 (2.6%) as severe, respectively.

At the time of the follow-up interview, 1115 (42.1%) of the participants reported no symptoms, 444 (16.8%) reported one, 313 (11.8%) two and 777 (29.3%) three or more symptoms, with fatigue, shortness of breath, and forgetfulness being the most common. Just under half (1247;47.1%) reported one or more LS. Persistent fatigue 551/2599 (21.2%), breathlessness 378/2614 (14.5%), forgetfulness 237/2597 (9.1%), muscle weakness 199/2592 (7.7%), problems seeing 198/2598 (7.6%), hair loss 183/2580 (7.1%), and problems sleeping 180/2583 (7%) were the most common LS reported at follow-up. Detailed information on all the symptoms, including duration, is presented in **Table S2**.

Although many patients had LS since discharge, some participants reported at least one symptom of a differing duration during follow-up interview; 285 (10.8%) had experienced these symptoms for 3 to 6 months, 179 (6.8%) between 2 and 3 months, 157 (5.9%) between 1 and 2 months, 103 (3.9%) between 2 and 4 weeks and 140 (5.3%) between 1 and 2 weeks, respectively. The duration of the ten common symptoms at the time of the follow-up is shown in **Figure 2**.

**Figure 2.**
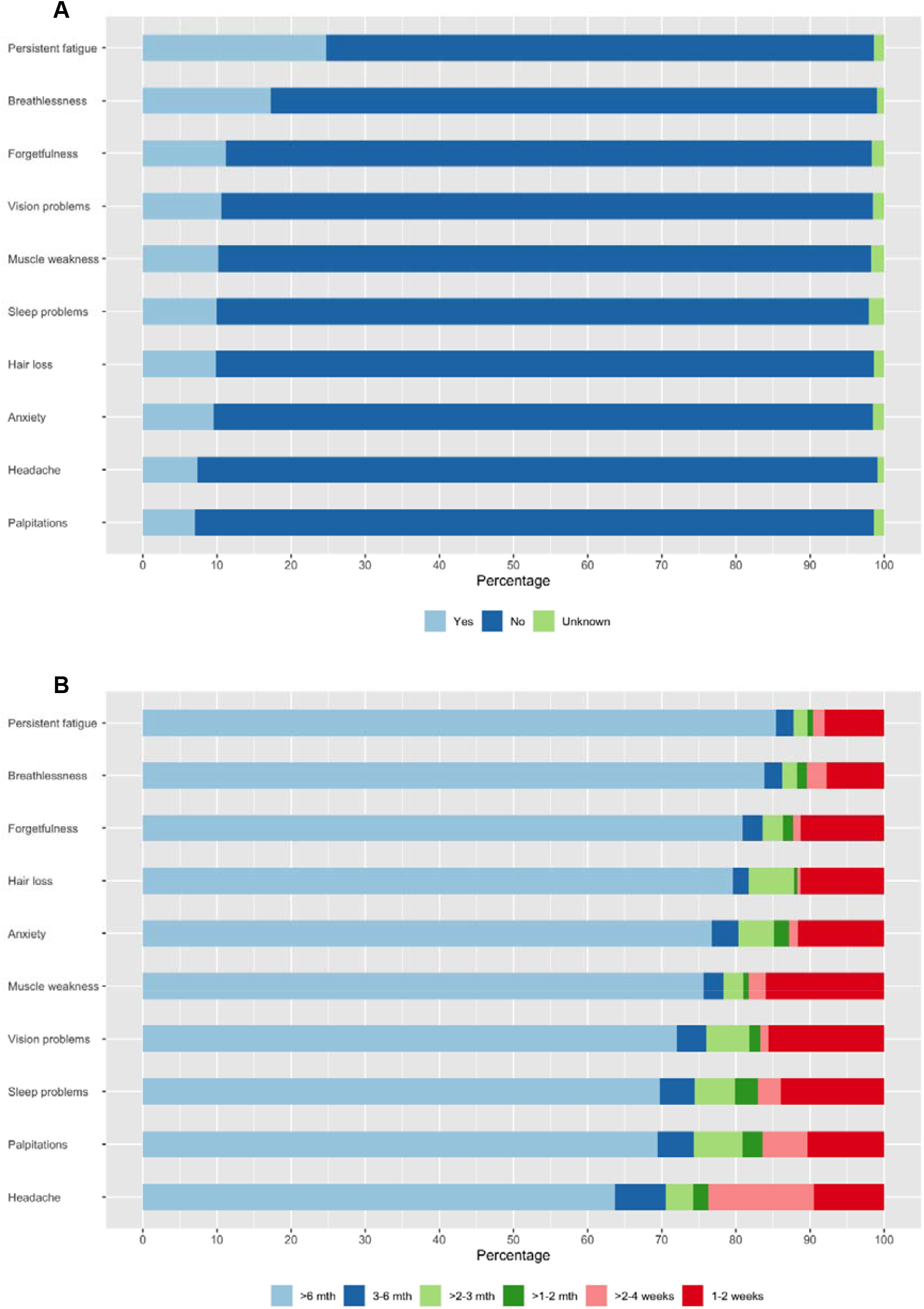
Stacked bar charts presenting (a) prevalence and (b) duration of ten most common symptoms reported at the time of the follow-up interview. The full data is shown in the Table S2.

A degree of overlap was found between the five most common LS, with 79/900 (8.8%) of patients experiencing both persistent fatigue and breathlessness, 54 (6%) persistent fatigue and muscle weakness. A smaller proportion of patients reported a combination of persistent fatigue, breathlessness and muscle weakness - 26/900 (2.9%) with 16 (1.8%) patients having all five (**Figure S1**).

With regard to categories of LS, chronic fatigue was found to be the most common 658/2593 (25%) at the time of the follow-up interview, followed by respiratory 451/2616 (17.2%), neurological 375/2586 (14.5%), mood and behaviour changes 284/2591 (11%) and dermatological 206/2583 (8%) symptoms. A smaller number of patients experienced gastrointestinal 110/2599 (4.2%) and sensory 70/2622 (2.7%) problems since discharge.

A small number of the LS categories were co-existent: 174 (6.6%) participants reported LS from three different categories at the time of the follow-up interview;88 (3.3%) reported four categories, and 37 (1.4%) reported five categories or more. Co-existence of five most common categories of long-standing symptoms at the time of the follow-up interview is presented in the **Figure S1**.

Risk factors for all categories were assessed. In multivariable regression analysis, female sex was a predictor of chronic fatigue with an odds ratio of 1.67 (95% confidence interval 1.39 to 2.02), neurological (2.03, 1.60 to 2.58), mood and behaviour (1.83, 1.41 to 2.40), dermatological (3.26, 2.36 to 4.57), gastrointestinal (2.50, 1.64 to 3.89), sensory (1.73, 2.06 to 2.89) and respiratory (1.31, 1.06 to 1.62) long-standing symptom categories, respectively. The effect of female sex remained unchanged in the sensitivity analyses, which included patients with RT-PCR-confirmed COVID-19 only, for all categories except respiratory and sensory. Pre-existing asthma was consistently associated with neurological (1.95, 1.25 to 2.98) and mood and behavioural changes (2.02, 1.24 to 3.18) (**Figures 3 and 4**), with associations remaining significant in the sensitivity analyses. Chronic pulmonary disease was associated with chronic fatigue (1.68, 1.21 to 2.32) (**Figure 5**) and gastrointestinal (1.93, 1.02 to 3.43) long-standing symptom categories development. However, an association with gastrointestinal symptoms was not confirmed in the sensitivity analysis. Rheumatological disorder was associated with the mood and behavioural long-standing symptoms (1.97, 1.12-3.33), but the effect was not confirmed in the sensitivity analysis. Confirmed RT-PCR during acute phase was significantly associated with chronic fatigue, neurological, mood and behaviour and gastrointestinal categories, confirming importance of the sensitivity analyses. More details of primary and sensitivity analyses are presented in **Table 2** and forest plots are available as **supplementary material**.

**Table 2.** Risk factors significantly associated with the different categories of long-standing symptoms in the primary (age, sex, comorbidities, severity and RT-PCR were included as potential risk factors) and sensitivity (performed in a subgroup of RT-PCR positive patients only) multivariable regression analyses. *- The assessment of robustness is based on the magnitude, direction and/or statistical significance of the estimates. †- Number of patients with at least one long-standing symptom from this category) Statistically significant associations are presented in bold. Abbreviations: OR, Odds Ratio; CI, Confidence interval; NA, Not applicable; RT-PCR “+”, Real-time polymerase chain reaction confirmed SARS-CoV-2 infection.

| Long-standing symptom Category (n<sup>†</sup>) | Risk factor | Primary analysis OR (95%CI) | Sensitivity analysis OR (95%CI) | Consistently associated factors* |
| --- | --- | --- | --- | --- |
| Chronic fatigue (n = 658) | Female Sex | **1.67 (1.39-2.02)** | **1.81 (1.40-2.34)** | Female Sex |
|  | Chronic pulmonary disease | **1.68 (1.21-2.32)** | **1.66 (2.08-2.54)** | Chronic pulmonary disease |
|  | Hypertension | **1.27 (1.02-1.57)** | 1.27 (0.94-1.72) |  |
|  | RT-PCR “+” | **1.23 (1.02-1.47)** | NA |  |
| Respiratory (n = 451) | Female Sex | **1.31 (1.06-1.62)** | 1.21 (0.90-1.62) | No |
| Neurological (n = 375) | Female Sex | **2.03 (1.60-2.58)** | **2.21 (1.54-2.93)** | Female Sex |
|  | Asthma | **1.95 (1.25-2.98)** | **2.18 (1.24-3.73)** | Asthma |
|  | RT-PCR “+” | **1.30 (1.04-1.63)** | NA |  |
| Mood and behaviour (n = 284) | Female Sex | **1.83 (1.41-2.40)** | **1.73 (1.22-2.47)** | Female Sex |
|  | Asthma | **2.02 (1.24-3.18)** | **2.39 (1.31-4.20)** | Asthma |
|  | Rheumatological disorder | **1.97 (1.12-3.33)** | 1.62 (0.69-3.45) |  |
|  | RT-PCR “+” | **1.45 (1.12-1.87)** | NA |  |
| Gastrointestinal (n = 206) | Female Sex | **2.50 (1.64-3.89)** | **2.78 (1.61-4.96)** | Female Sex |
|  | Chronic pulmonary disease | **1.93 (1.02-3.43)** | 1.36 (0.54-2.99) |  |
|  | RT-PCR “+” | **1.56 (1.05-2.33)** | NA |  |
| Dermatological (n = 110) | Female Sex | **3.26 (2.36-4.57)** | **3.08 (1.99-4.88)** | Female Sex |
| Sensory (n = 70) | Female Sex | **1.73 (2.06-2.89)** | 1.54 (0.78-3.07) | No |

**Figure 3.**
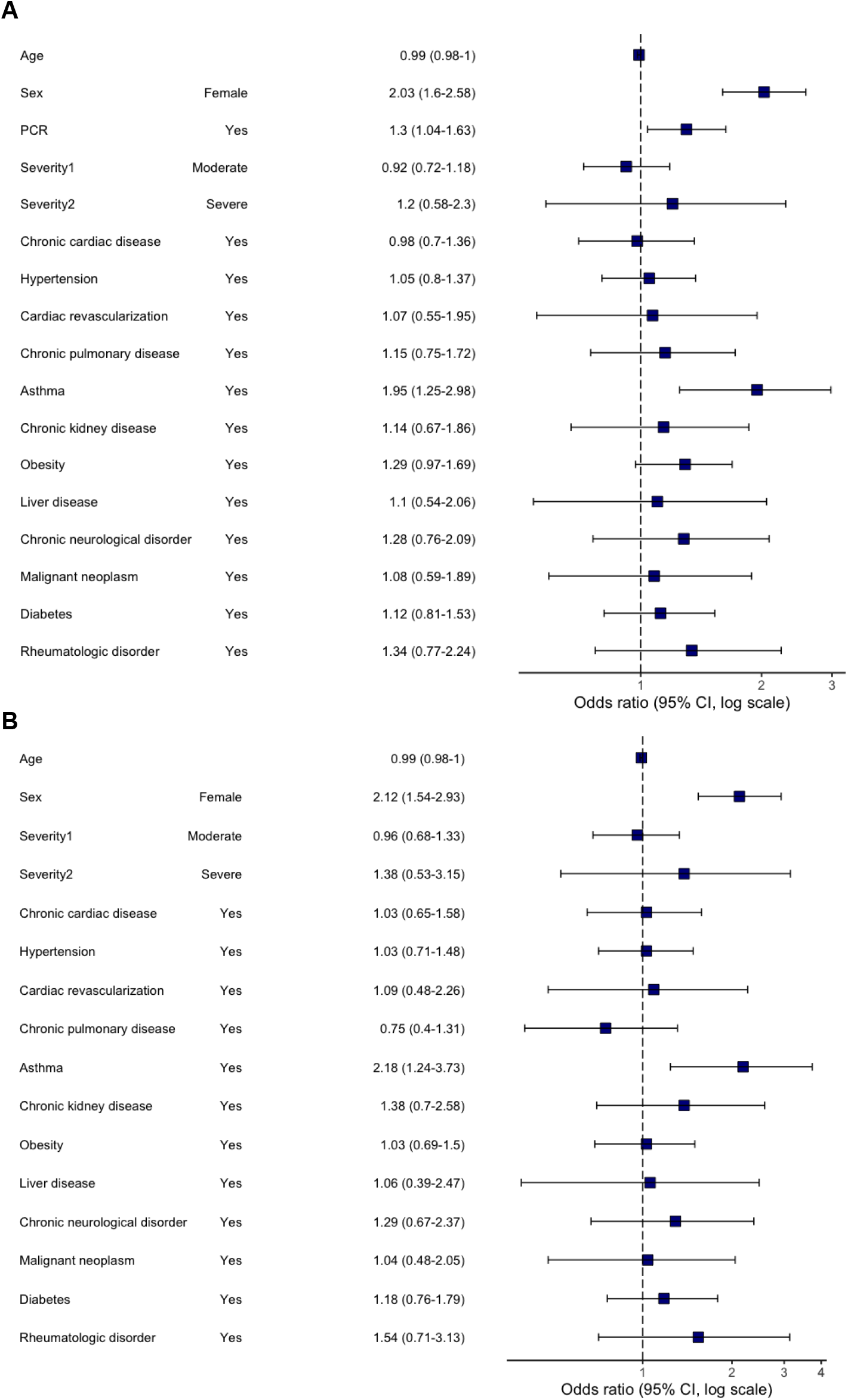
Multivariable logistic regression model. Odds ratios and 95% CIs for “Neurological” category of long-standing symptoms at the time of follow-up. Abbreviation: CI, confidence interval. (a) primary analysis (age, sex, comorbidities, severity and RT-PCR were included as potential risk factors); (b) sensitivity analysis (performed in a subgroup of RT-PCR positive patients only).

**Figure 4.**
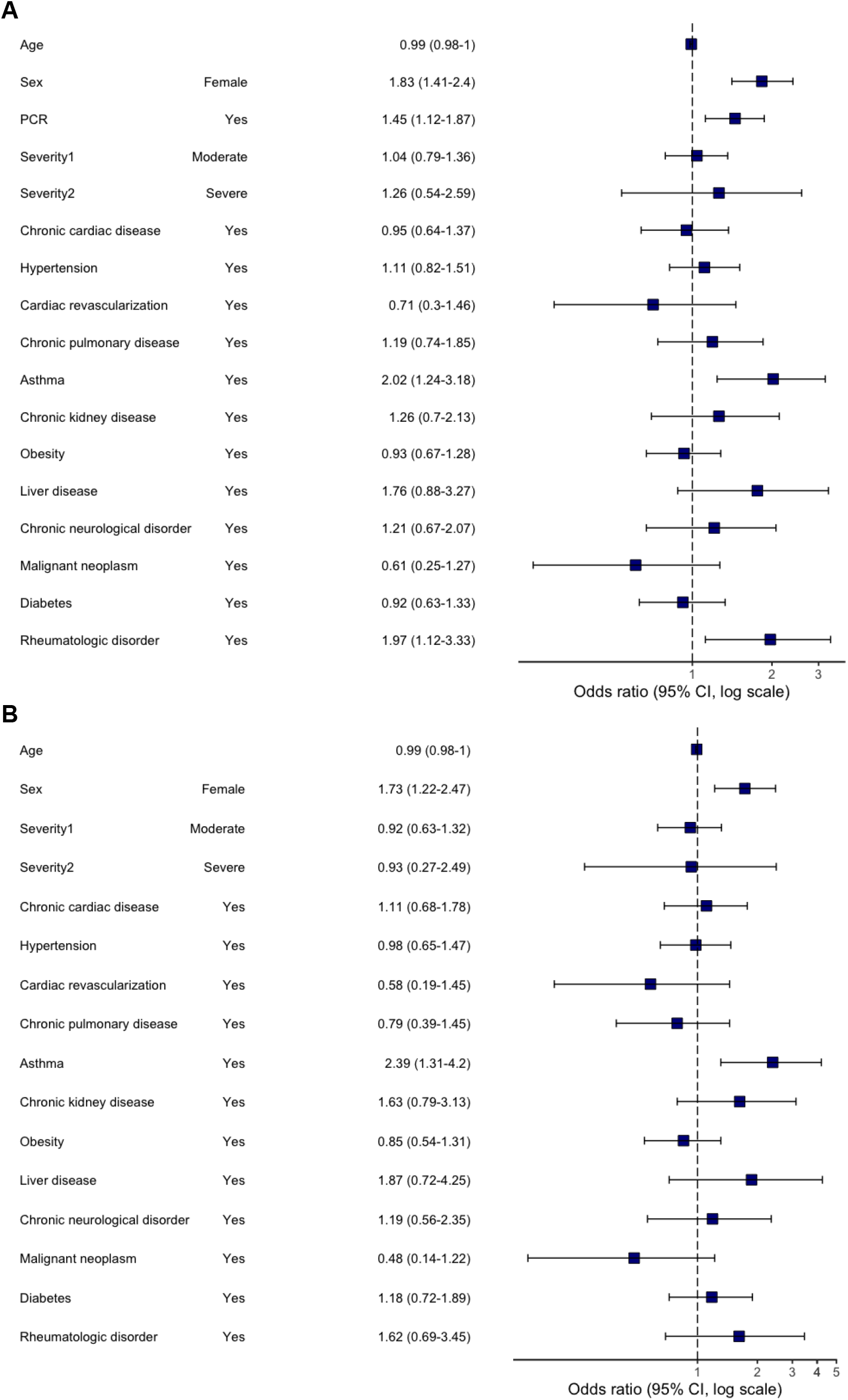
Multivariable logistic regression model. Odds ratios and 95% CIs for “Mood and behaviour” category of long-standing symptoms at the time of follow-up. Abbreviation: CI, confidence interval. (a) primary analysis (age, sex, comorbidities, severity and RT-PCR were included as potential risk factors); (b) sensitivity analysis (performed in a subgroup of RT-PCR positive patients only).

**Figure 5.**
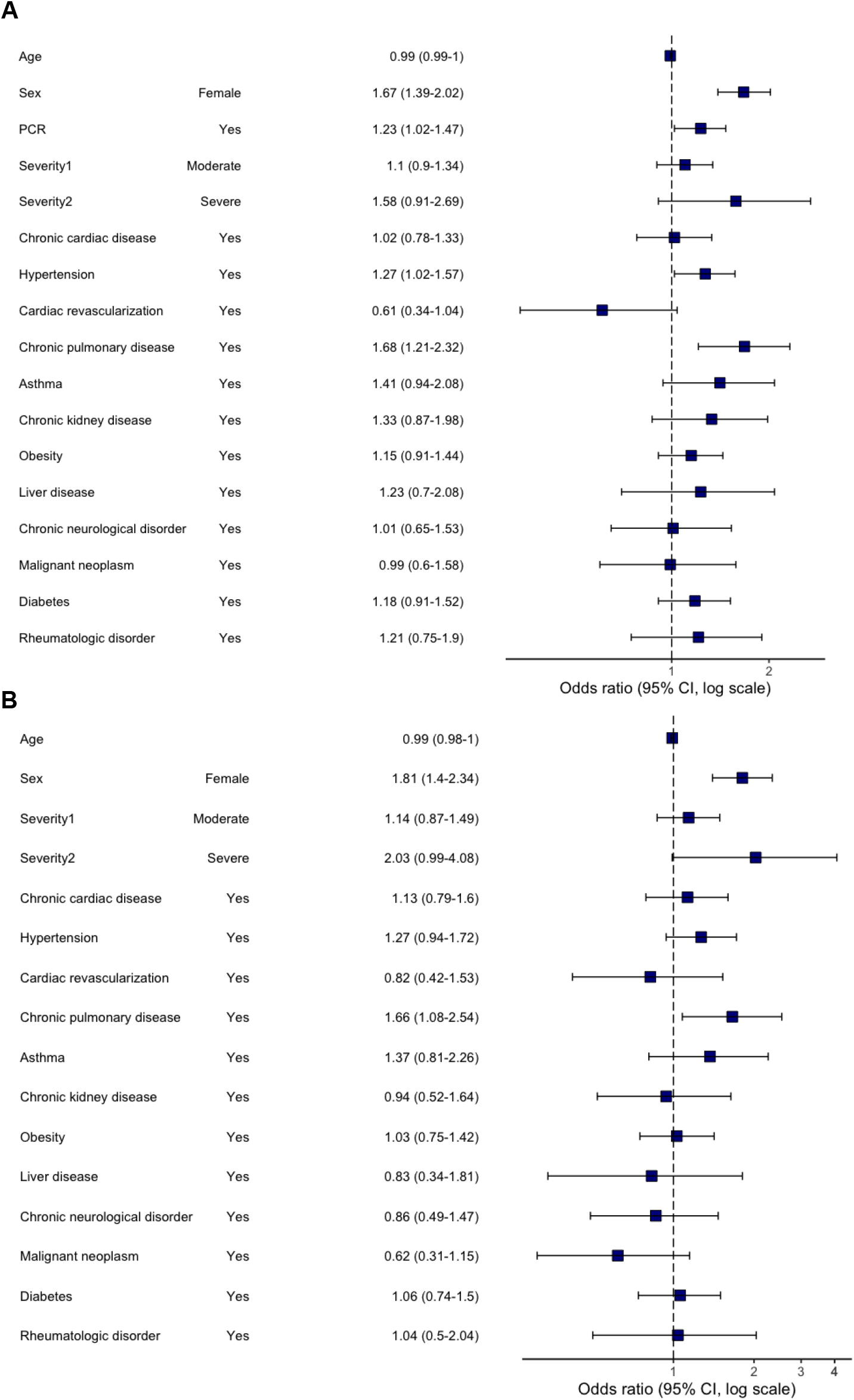
Multivariable logistic regression model. Odds ratios and 95% CIs for “Chronic fatigue” category of long-standing symptoms at the time of follow-up. Abbreviation: CI, confidence interval. (a) primary analysis (age, sex, comorbidities, severity and RT-PCR were included as potential risk factors); (b) sensitivity analysis (performed in a subgroup of RT-PCR positive patients only)

Dyspnoea of different severity was reported by 318 (12%) patients during follow-up with 194 (7.3%) equivalent to grade 3, 93 (3.5%) grade 4 and 31 (1.2%) grade 5 according to MRC Dyspnoea Scale (**Table S3**).

Participants reported lower scores (poorer health state) on the EuroQol visual analog scale at follow-up compared with pre-COVID-19 onset, median 80 (IQR, 65-90) vs 85 (70-95) (p<0.001). Significant worsening of the health state compared with pre-COVID 19 was found across all symptom categories, with the highest median difference reported by patients with gastrointestinal (−15), mood and behaviour (−13) and neurological (−10.5) symptoms (p<0.001 for all) (**Table S4**). No statistically significant reduction in health state was found among patients reporting no symptoms. Participants falling into all symptom categories had significantly lower health state than those with no symptoms (p<0.001 for all).

## DISCUSSION

To our knowledge, this is the prospective cohort study with the largest sample size and the longest follow-up duration, assessing the long-term health and psycho-social consequences of COVID-19 in hospitalised adults. The cohort included a similar number of RT-PCR-confirmed COVID-19 and those who were clinically diagnosed with COVID-19. We found that six of ten patients experienced at least one symptom of any duration 6 to 8 months after hospital discharge and almost a half of the patients reported at least one LS, with chronic fatigue and respiratory problems being the most frequent LS categories. One in ten patients reported multisystem impacts with three or more categories of long-standing symptoms present at follow-up. LS were experienced by males and females, with a higher risk amongst women, with chronic pulmonary disease associated with chronic fatigue, and asthma with a higher risk of neurological symptoms and mood and behaviour problems.

The majority of patients experienced LS from the time of discharge, with a smaller number developing symptoms months following discharge. Persistent fatigue and breathlessness were the most frequent LS in our cohort, which is consistent with recent report from China ^6^. Forgetfulness and vision problems were also among the most common, while problems sleeping was less common (10.2%) compared to rates reported by the recent follow-up data from China (26%) ^6^.

A novel findings relates to the development of symptoms weeks or months since recovery from COVID-19. To our knowledge, this aspect has not been investigated in previous studies, as most of the cohorts did not collect data on the duration of the symptoms present at follow-up. Patterns of the symptom development following COVID-19 should be further investigated in future research.

Female sex was significantly associated with an increased risk of LS, regardless of symptom category, reflecting previous findings ^6^ and digital App ^16^ studies. Chronic pulmonary disease was a risk factor for the development of chronic fatigue. An association between chronic pulmonary disease and severe acute COVID-19 was found in many studies ^17^, but it has not been previously reported as a risk factor for COVID-19 sequelae. The presence of chronic pulmonary disease has been previously associated with chronic fatigue syndrome ^18^. The pandemic also had a significant adverse impact on care and support for patients with chronic pulmonary conditions, including a reduction in face-to-face clinic availability, lack of access to pulmonary rehabilitation sessions and hospital care during an exacerbation due to fear of COVID-19 exposure ^19^. The causality cannot be determined and we are unable to conclude if lack of follow-up and involvement in rehabilitation programmes for chronic pulmonary conditions was the cause of ongoing symptoms. Future research should investigate COVID-19 consequences in this group of patients in greater detail.

Data from the COVID Symptom Study app in the UK suggested that asthma may be a risk factor for LS related to COVID-19 ^16^. We also found that asthma was associated with an increased risk of LS during follow-up, specifically neurological and mood and behaviour. Although asthma has not been associated with a higher risk of hospital admission and/or in-hospital mortality in COVID-19 patients, ^20,21^ different results may be found when considering the long-term consequences of infection. Recent research suggested that COVID-19 sequelae may be associated with the mast cell activation syndrome ^22^ and the Th-2 biased immunological response in asthmatic patients may be responsible for an increased risk of long-term implications of the infection. This finding may point to immune mediated mechanisms but requires confirmation in a larger sample size with a more detailed investigation, including in-clinic visits.

It is worth noting that in contrast to the Chinese cohort ^6^, we did not find significant associations between COVID-19 severity and long-term symptom development. This may be, however, related to the difference in the methodology applied and the small number of severe patients in our cohort. There is a need for further investigation of the frequency with which mild infection are followed by long-term symptoms, mandating a study of non-hospitalised infected subjects.

Patients with all categories of LS reported significantly lower health state when compared with symptom-free patients. They also considered the health state to be lower than before the COVID-19 episode. This is consistent with previous reports from different countries ^5,6,23^. This finding points to the multi-factorial adverse effects of COVID-19 and to the need for wide ranging and longer term supports.

A major strength of this study is use of pre-positioned data collection method using ISARIC Core CRF for acute phase data and ISARIC Long-term Follow-up Study CRF. Another strength is the large sample size, as this cohort is the largest follow-up assessment of hospitalised adults to date. Stratification to determine if the symptoms were persistent following COVID-19 was another novel aspect of the study. At the same time, this cohort study has some limitations. First, the study population only included patients within Moscow, although regional clustering is common to all major cohort studies published during the COVID-19 pandemic. Second, acute data were collected from the electronic medical records with no access to additional information that could be potentially retrieved from the medical notes. Third, almost half of the patients in our cohort did not have RT-PCR confirmed COVID-19 infection, although our previous work 3 showed that clinical features of COVID-19 and in-hospital mortality were the same in COVID-19 clinically diagnosed and laboratory-confirmed cases. We also performed sensitivity analyses using data from the laboratory-confirmed COVID-19 patients only to ensure consistency and robustness of the findings. Fifth, some patients may have developed additional comorbidities or complications since the hospital discharge, which were not appropriately captured and could potentially affect the QoL and symptom prevalence and persistence. There is also a risk of recall bias in reporting quality of life and dyspnoea preceding COVID-19.

At 6-8 months follow-up, many patients experienced symptoms from the time of hospital discharge, with chronic fatigue and respiratory problems being the most common sequelae. Most patients reported symptoms at 6-8 months from the time of discharge, although a subgroup reported symptoms limited to a few weeks and/or months after the acute phase. One in ten individuals had multi-system involvement at the time of the follow-up. Female sex was the main risk factor for most of the long-lasting symptom categories development, while chronic pulmonary disease was associated with a higher risk of chronic fatigue development and asthma with the neurological and mood and behaviour changes. Future studies should focus on patients with multisystem involvement and longer follow-up of a large sample will allow for a better understanding of COVID-19 sequelae and help with the phenotype recognition. Investigation of immunological aspects of the association between asthma and several long-covid outcomes may identify mechanisms and therapeutic targets for therapy to mitigate adverse consequences.

## Data Availability

The data that support the findings of this study are available from the corresponding author, DM, upon reasonable request and Institutional approval.

## Acknowledgments

We thank RFBR, grant 20-04-60063 for supporting the work. We would also like to thank UK Embassy in Moscow for providing a grant supporting our project. We are very grateful to the Sechenov University Hospital Network clinical staff and to the patients, carers and families for their kindness and understanding during these difficult times of COVID-19 pandemic. We would like to express our very great appreciation to ISARIC Global COVID-19 follow-up working group for the survey development. We would like to thank Mr Maksim Kholopov for providing technical support in data collection and database administration. We are very thankful to Eat & Talk, Luch, Black Market and Academia for providing us the work space in time of need. We are grateful to Ms Asmik Avagyan, Ms Daria Belykh, Ms Ekaterina Belyakova, Ms Anna Berbenyuk, Mr Dmitry Eliseev, Ms Mariia Grosheva, Ms Nelli Khusainova, Ms Maria Kislova, Ms Valeria Klishina, Ms Karina Kovygina, Ms Natalia Kogut, Ms Yana Kohanovskaya, Ms Anastasia Kuznetsova, Ms Elza Lidzhieva, Ms Nadezhda Markina, Mr Georgiy Novoselov, Ms Anna Pushkareva, Ms Olga Romanova, Ms Maria Shoshorina, Ms Jasmin Sibkhan, Ms Olga Spasskaya, Ms Anna Surkova, Ms Nailya Urmantaeva, Ms Ekaterina Varlamova, Ms Margarita Yegiyan, Ms Margarita Zaikina, Ms Anastasia Zorina, Ms Elena Zuikova for assistance in data extraction, document translation and help during the project. Finally, we would like to extend our gratitude to the Global ISARIC team and ISARIC Co0ordinating Centre for their continuous support, expertise and for the development of the outbreak ready standardised protocols for the data collection.

