## Supplementary material for "Risk factors for long-term consequences of COVID-19 in hospitalised adults in Moscow using the ISARIC Global follow-up protocol: StopCOVID cohort study"

### **Case Report Form (CRF) of Initial Survey: First Follow-Up Time Point^[1]^**

**1.** **About you and your COVID-19 illness (if you’re completing this survey on behalf of**

**a child or adult that you care for, all the questions relate to their health and wellbeing)**

Date you did the survey (DD/MM/YYYY):	[_D_][_D_]/[_M_][_M_]/[_2_][_0_][_Y_][_Y_]

What is your date of birth (DD/MM/YYYY):	[_D_][_D_]/[_M_][_M_]/[_Y_][_Y_][_Y_][_Y_]

Have you been vaccinated against influenza within last 6 months?	 Yes	 No	 Not sure

Have you had a pneumococcal vaccination within the last 5 years?	 Yes	 No	 Not sure

Please grade the severity of your COVID-19 illness:  Mild  Moderate  Severe  Critical

Roughly what day did you first

experience symptoms of COVID-19?	[_D_][_D_]/[_M_][_M_]/[_2_][_0_][_Y_][_Y_]

Were you admitted to hospital due to COVID-19?	 Yes	 No

- Roughly at what date were you first

admitted to hospital?	[_D_][_D_]/[_M_][_M_]/[_2_][_0_][_Y_][_Y_]

- Roughly at what date were you first

discharged from hospital?	[_D_][_D_]/[_M_][_M_]/[_2_][_0_][_Y_][_Y_]

- Have you been re-admitted to hospital or

health facility after your Covid-19 illness?	 Yes	 No

If yes, how many times: [_Number_]

If yes, specify reason:

Name of hospital/s:

Did you have a positive antibody test ? ☐ Yes ☐ No ☐ Not sure

If yes, please confirm the date of positive antibody test: [_D_][_D_]/[_M_][_M_]/[_Y_][_Y_][_Y_][_Y_]

If ever admitted to hospital/health facility for Covid-19,

were you admitted to intensive care (ICU/ITU)?  Yes	 No	 Not sure

**2. About your health now**

**Do you feel fully recovered from COVID-19?**

 Strongly disagree  Disagree  Slightly disagree  Slightly agree  Agree  Strongly Agree

**Have you felt feverish recently?**  Yes  No  Not sure

If yes roughly when did	 within last 7 days	 between 1 to 2 weeks ago you last feel feverish?	 between 2 to 4 weeks ago	 between 1 to 2 months ago

 between 2 to 3 months ago

**If yes, what was the cause**  COVID -19	 Other respiratory infection (cough/cold/sore throat) **of your most recent**	 Stomach infection (diarrhoea/vomiting)	 Urinary infection **feverish illness?**	 TB	 Other: specify:

 Unknown  Prefer not to say

**3. Since having COVID-19, have you been diagnosed with any of these?**

Heart attack	 Yes  No	Deep vein thrombosis (DVT, “Clot in leg”)  Yes  No

Stroke or mini stroke/TIA	 Yes  No	Pulmonary embolism (PE, “Clot in lung”)  Yes  No

Kidney problems	 Yes  No	Other condition (please specify)?__________________

| **4. Within the last seven days, have you had any of these symptoms?** | | |  |  |
| --- | --- | --- | --- | --- |
| **Symptoms** | **Yes/No** | **If yes**, what is the duration of symptoms |  |  |
| Shortness of breath/breathlessness | □ Yes □ No | □ 1-2 weeks □ > 2-4 weeks □ >1-2 months □ >2-3 months □ >3-6 months □ from the time of discharge |  |  |
| Pain on breathing | □ Yes □ No | □ 1-2 weeks □ > 2-4 weeks □ >1-2 months □ >2-3 months □ >3-6 months □ from the time of discharge |  |  |
| Persistent cough | □ Yes □ No | □ 1-2 weeks □ > 2-4 weeks □ >1-2 months □ >2-3 months □ >3-6 months □ from the time of discharge |  |  |
| *If yes, □ dry cough □ with phlegm* | | |  |  |
| Weakness in arms or legs/ muscle weakness | □ Yes □ No | □ 1-2 weeks □ > 2-4 weeks □ >1-2 months □ >2-3 months □ >3-6 months □ from the time of discharge |  |  |
| Joint pain or swelling | □ Yes □ No | □ 1-2 weeks □ > 2-4 weeks □ >1-2 months □ >2-3 months □ >3-6 months □ from the time of discharge |  |  |
| Persistent muscle pain | □ Yes □ No | □ 1-2 weeks □ > 2-4 weeks □ >1-2 months □ >2-3 months □ >3-6 months □ from the time of discharge |  |  |
| Headache | □ Yes □ No | □ 1-2 weeks □ > 2-4 weeks □ >1-2 months □ >2-3 months □ >3-6 months □ from the time of discharge |  |  |
| Dizziness/ light headedness | □ Yes □ No | □ 1-2 weeks □ > 2-4 weeks □ >1-2 months □ >2-3 months □ >3-6 months □ from the time of discharge |  |  |
| Fainting/ blackouts | □ Yes □ No | □ 1-2 weeks □ > 2-4 weeks □ >1-2 months □ >2-3 months □ >3-6 months □ from the time of discharge |  |  |
| Can't feel one side of the body or face | □ Yes □ No | □ 1-2 weeks □ > 2-4 weeks □ >1-2 months □ >2-3 months □ >3-6 months □ from the time of discharge |  |  |
| Loss of smell | □ Yes □ No | □ 1-2 weeks □ > 2-4 weeks □ >1-2 months □ >2-3 months □ >3-6 months □ from the time of discharge |  |  |
| Loss of taste | □ Yes □ No | □ 1-2 weeks □ > 2-4 weeks □ >1-2 months □ >2-3 months □ >3-6 months □ from the time of discharge |  |  |
| Myoclonus | □ Yes □ No | □ 1-2 weeks □ > 2-4 weeks □ >1-2 months □ >2-3 months □ >3-6 months □ from the time of discharge |  |  |
| Tremor/shakiness | □ Yes □ No | □ 1-2 weeks □ > 2-4 weeks □ >1-2 months □ >2-3 months □ >3-6 months □ from the time of discharge |  |  |
| Tingling feeling/ “pins and needles“ | □ Yes □ No | □ 1-2 weeks □ > 2-4 weeks □ >1-2 months □ >2-3 months □ >3-6 months □ from the time of discharge |  |  |
| Seizures/fits | □ Yes □ No | □ 1-2 weeks □ > 2-4 weeks □ >1-2 months □ >2-3 months □ >3-6 months □ from the time of discharge |  |  |
| Problems swallowing or chewing | □ Yes □ No | □ 1-2 weeks □ > 2-4 weeks □ >1-2 months □ >2-3 months □ >3-6 months □ from the time of discharge |  |  |
| Problems speaking or communicating | □ Yes □ No | □ 1-2 weeks □ > 2-4 weeks □ >1-2 months □ >2-3 months □ >3-6 months □ from the time of discharge |  |  |
| Problems with balance | □ Yes □ No | □ 1-2 weeks □ > 2-4 weeks □ >1-2 months □ >2-3 months □ >3-6 months □ from the time of discharge |  |  |
| Can’t fully move or control movement | □ Yes □ No | □ 1-2 weeks □ > 2-4 weeks □ >1-2 months □ >2-3 months □ >3-6 months □ from the time of discharge |  |  |
| Stiffness of muscles, rigidity | □ Yes □ No | □ 1-2 weeks □ > 2-4 weeks □ >1-2 months □ >2-3 months □ >3-6 months □ from the time of discharge |  |  |
| Slowness of movement (bradykinesia) | □ Yes □ No | □ 1-2 weeks □ > 2-4 weeks □ >1-2 months □ >2-3 months □ >3-6 months □ from the time of discharge |  |  |
| Double vision | □ Yes □ No | □ 1-2 weeks □ > 2-4 weeks □ >1-2 months □ >2-3 months □ >3-6 months □ from the time of discharge |  |  |
| Problems seeing | □ Yes □ No | □ 1-2 weeks □ > 2-4 weeks □ >1-2 months □ >2-3 months □ >3-6 months □ from the time of discharge |  |  |
| Problems hearing | □ Yes □ No | □ 1-2 weeks □ > 2-4 weeks □ >1-2 months □ >2-3 months □ >3-6 months □ from the time of discharge |  |  |
| Confusion/lack of concentration | □ Yes □ No | □ 1-2 weeks □ > 2-4 weeks □ >1-2 months □ >2-3 months □ >3-6 months □ from the time of discharge |  |  |
| Hallucinations (seeing or hearing things others don’t see or hear) | □ Yes □ No | □ 1-2 weeks □ > 2-4 weeks □ >1-2 months □ >2-3 months □ >3-6 months □ from the time of discharge |  |  |
| Problems sleeping | □ Yes □ No | □ 1-2 weeks □ > 2-4 weeks □ >1-2 months □ >2-3 months □ >3-6 months □ from the time of discharge |  |  |
| Anxiety | □ Yes □ No | □ 1-2 weeks □ > 2-4 weeks □ >1-2 months □ >2-3 months □ >3-6 months □ from the time of discharge |  |  |
| Forgetfulness | □ Yes □ No | □ 1-2 weeks □ > 2-4 weeks □ >1-2 months □ >2-3 months □ >3-6 months □ from the time of discharge |  |  |
| Loss of interest or pleasure | □ Yes □ No | □ 1-2 weeks □ > 2-4 weeks □ >1-2 months □ >2-3 months □ >3-6 months □ from the time of discharge |  |  |
| Behavioural changes | □ Yes □ No | □ 1-2 weeks □ > 2-4 weeks □ >1-2 months □ >2-3 months □ >3-6 months □ from the time of discharge |  |  |
| Depressed mood | □ Yes □ No | □ 1-2 weeks □ > 2-4 weeks □ >1-2 months □ >2-3 months □ >3-6 months □ from the time of discharge |  |  |
| Persistent fatigue | □ Yes □ No | □ 1-2 weeks □ > 2-4 weeks □ >1-2 months □ >2-3 months □ >3-6 months □ from the time of discharge |  |  |
| Diarrhea | □ Yes □ No | □ 1-2 weeks □ > 2-4 weeks □ >1-2 months □ >2-3 months □ >3-6 months □ from the time of discharge |  |  |
| Stomach/ abdominal pain | □ Yes □ No | □ 1-2 weeks □ > 2-4 weeks □ >1-2 months □ >2-3 months □ >3-6 months □ from the time of discharge |  |  |
| Feeling sick | □ Yes □ No | □ 1-2 weeks □ > 2-4 weeks □ >1-2 months □ >2-3 months □ >3-6 months □ from the time of discharge |  |  |
| Vomiting | □ Yes □ No | □ 1-2 weeks □ > 2-4 weeks □ >1-2 months □ >2-3 months □ >3-6 months □ from the time of discharge |  |  |
| Constipation | □ Yes □ No | □ 1-2 weeks □ > 2-4 weeks □ >1-2 months □ >2-3 months □ >3-6 months □ from the time of discharge |  |  |
| Palpitations (heart racing) | □ Yes □ No | □ 1-2 weeks □ > 2-4 weeks □ >1-2 months □ >2-3 months □ >3-6 months □ from the time of discharge |  |  |
| Swollen ankle(s) | □ Yes □ No | □ 1-2 weeks □ > 2-4 weeks □ >1-2 months □ >2-3 months □ >3-6 months □ from the time of discharge |  |  |
| ~~Erectile dysfunction~~ | ~~□ Yes □ No~~ | ~~□ 1-2 weeks □ > 2-4 weeks □ >1-2 months □ >2-3 months □ >3-6 months □ from the time of discharge~~ |  |  |
| Problems passing urine | □ Yes □ No | □ 1-2 weeks □ > 2-4 weeks □ >1-2 months □ >2-3 months □ >3-6 months □ from the time of discharge |  | **If yes**, what is the duration of symptoms |
| Changes in menstruation | □ Yes □ No | □ 1-2 weeks □ > 2-4 weeks □ >1-2 months □ >2-3 months □ >3-6 months □ from the time of discharge |  |  |
| Lumps or rashes (purple/pink) on toes | □ Yes □ No | □ 1-2 weeks □ > 2-4 weeks □ >1-2 months □ >2-3 months □ >3-6 months □ from the time of discharge |  |  |
| Hair loss | □ Yes □ No | □ 1-2 weeks □ > 2-4 weeks □ >1-2 months □ >2-3 months □ >3-6 months □ from the time of discharge |  |  |
| Skin rash  *If yes, tick all body areas that apply:* | □ Yes □ No | □ 1-2 weeks □ > 2-4 weeks □ >1-2 months □ >2-3 months □ >3-6 months □ from the time of discharge |  |  |
| *Face* | □ Yes □ No | □ 1-2 weeks □ > 2-4 weeks □ >1-2 months □ >2-3 months □ >3-6 months □ from the time of discharge |  |  |
| *Trunk (stomach or back)* | □ Yes □ No | □ 1-2 weeks □ > 2-4 weeks □ >1-2 months □ >2-3 months □ >3-6 months □ from the time of discharge |  |  |
| *Arms* | □ Yes □ No | □ 1-2 weeks □ > 2-4 weeks □ >1-2 months □ >2-3 months □ >3-6 months □ from the time of discharge |  |  |
| *Legs* | □ Yes □ No | □ 1-2 weeks □ > 2-4 weeks □ >1-2 months □ >2-3 months □ >3-6 months □ from the time of discharge |  |  |
| *Buttocks* | □ Yes □ No | □ 1-2 weeks □ > 2-4 weeks □ >1-2 months □ >2-3 months □ >3-6 months □ from the time of discharge |  |  |
| *Toes* | □ Yes □ No | □ 1-2 weeks □ > 2-4 weeks □ >1-2 months □ >2-3 months □ >3-6 months □ from the time of discharge |  |  |
| *Fingers* | □ Yes □ No | □ 1-2 weeks □ > 2-4 weeks □ >1-2 months □ >2-3 months □ >3-6 months □ from the time of discharge |  |  |
| *Accompanied by itch* | □ Yes □ No | □ 1-2 weeks □ > 2-4 weeks □ >1-2 months □ >2-3 months □ >3-6 months □ from the time of discharge |  |  |
| Chest pains | □ Yes □ No | □ 1-2 weeks □ > 2-4 weeks □ >1-2 months □ >2-3 months □ >3-6 months □ from the time of discharge |  |  |
| Hyperhidrosis | □ Yes □ No | □ 1-2 weeks □ > 2-4 weeks □ >1-2 months □ >2-3 months □ >3-6 months □ from the time of discharge |  |  |
| Bleeding | □ Yes □ No | □ 1-2 weeks □ > 2-4 weeks □ >1-2 months □ >2-3 months □ >3-6 months □ from the time of discharge |  |  |
| *If yes, specify bleeding site:* | □ Yes □ No |  |  |  |
| Weight loss | □ Yes □ No | □ 1-2 weeks □ > 2-4 weeks □ >1-2 months □ >2-3 months □ >3-6 months □ from the time of discharge |  |  |
| Loss of appetite | □ Yes □ No | □ 1-2 weeks □ > 2-4 weeks □ >1-2 months □ >2-3 months □ >3-6 months □ from the time of discharge |  |  |
| Any other NEW symptoms?  If yes, specify________________________ | □ Yes □ No | □ 1-2 weeks □ > 2-4 weeks □ >1-2 months □ >2-3 months □ >3-6 months □ from the time of discharge |  |  |

| **5. About your health** | | | | |
| --- | --- | --- | --- | --- |
| Under each heading, please tick the ONE box that describes your health BEFORE your COVID19 illness | | | | |
| **MOBILITY** | | **SELF-CARE** | | |
| I had no problems in walking about |  | I had no problems in walking about | |  |
| I had slight problems in walking about |  | I had slight problems in walking about | |  |
| I had moderate problems in walking about  | | I had moderate problems washing or dressing myself  | | |
| I had severe problems in walking about |  | I had severe problems in walking about | |  |
| I was unable to walk about |  | I was unable to walk about | |  |
| **USUAL ACTIVITIES**  (e.g. work, study, housework, family or leisure activities)  I had no problems doing my usual activities   I had slight problems doing my usual activities   I had moderate problems doing my usual activities   I had severe problems doing my usually activities   I was unable to do my usual activities  | | | **PAIN / DISCOMFORT**  I had no pain or discomfort   I had slight pain or discomfort   I had moderate pain or discomfort   I had severe pain or discomfort   I had extreme pain or discomfort  | |
| **ANXIETY/DEPRESSION**  I was not anxious or depressed  I was slightly anxious or depressed  I was moderately anxious or depressed I was severely anxious or depressed  I was extremely anxious or depressed |          |  |  |  |
| Under each heading, please tick the ONE box that best describes your health **TODAY** | | | | |
| **MOBILITY** | | **SELF-CARE**  I have no problems washing or dressing myself  I have slight problems washing or dressing myself   I have moderate problems washing or dressing myself   I have severe problems washing or dressing myself   I am unable to wash or dress myself  | | |
| I have no problems in walking about |  |  |  |  |
| I have slight problems in walking about |  |  |  |  |
| I have moderate problems in walking about  | |  |  |  |
| I have severe problems in walking about |  |  |  |  |
| I am unable to walk about |  |  |  |  |
| **USUAL ACTIVITIES**  (e.g. work, study, housework, family or leisure activities)  I have no problems doing my usual activities   I have slight problems doing my usual activities   I have moderate problems doing my usual activities   I have severe problems doing my usually activities   I am unable to do my usual activities  | | | **PAIN / DISCOMFORT**  I have no pain or discomfort  I have slight pain or discomfort   I have moderate pain or discomfort   I have severe pain or discomfort   I have extreme pain or discomfort  | |
| **ANXIETY/DEPRESSION**  I am not anxious or depressed  I am slightly anxious or depressed  I am moderately anxious or depressed I am severely anxious or depressed  I am extremely anxious or depressed |          |  |  | |

| 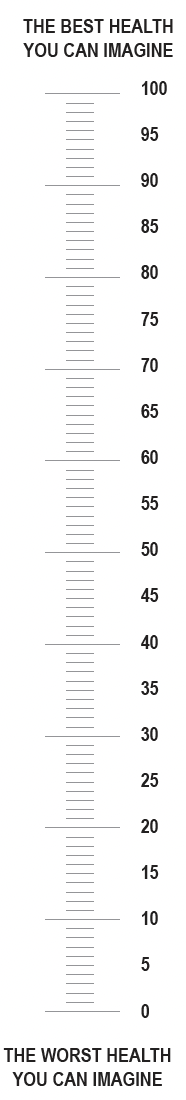   - We would like to know how good or bad your health   BEFORE COVID-19.     - This scale is numbered from 0 to 100. - 100 means the best health you can imagine.   0 means the worst health you can imagine.   - Mark an X on the scale to indicate how your health is   BEFORE COVID-19.   - Now, please write the number you marked on the   scale in the box below  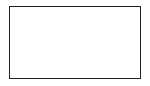  **YOUR HEALTH BEFORE COVID-19** =     - We would like to know how good or bad your health   TODAY.     - This scale is numbered from 0 to 100. - 100 means the best health you can imagine.   0 means the worst health you can imagine.   - Mark an X on the scale to indicate how your health is   TODAY.   - Now, please write the number you marked on the   scale in the box below  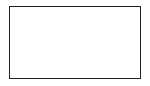  **YOUR HEALTH TODAY** = | | |
| --- | --- | --- |
| **6. Breathlessness and tiredness** | | |
| **Please tick ONE box that best describes how breathless you feel Today and ONE box that describes how BREATHLESS you felt before your Covid illness** | Within the last 24 hours  (tick one box) | Before your Covid19 illness (tick one box) |
| Not troubled by breathlessness except on strenuous exercise |  |  |
| Short of breath when hurrying or when walking up a slight hill |  |  |
| Walks slower than most people of my age because of breathlessness, or have to stop for breath when walking at own pace |  |  |
| Stops for breath after walking 100 yards/ 90-100 meters, or after a few minutes on level ground |  |  |
| Too breathless to leave the house, or breathless when dressing/undressing |  |  |
| **Please rate the intensity of your fatigue on average over the last 24 hours, on a scale from 0 – 10.**  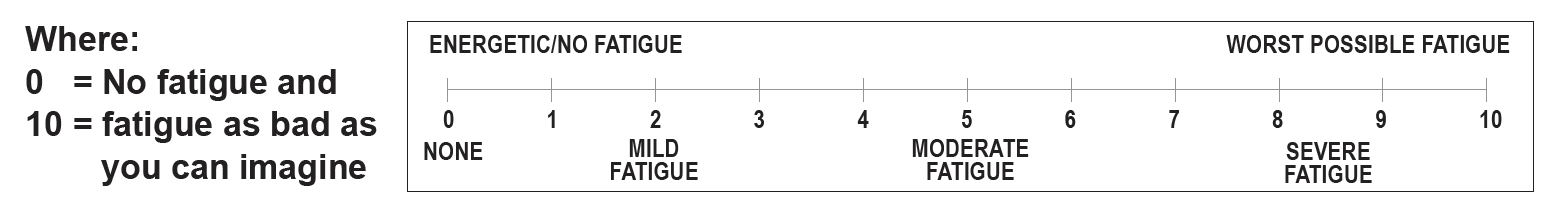 | | |

| **7. The next questions ask about difficulties you may have doing certain activities because of a HEALTH PROBLEM.** | | | | | | |
| --- | --- | --- | --- | --- | --- | --- |
| (mark the correct answer with a tick in the box) | | **Today** | | | **Before your Covid19 illness** | |
| Do you have difficulty seeing even if wearing glasses? | |  No - no difficulty   Yes – some difficulty   Yes – a lot of difficulty   Cannot do at all | | |  No - no difficulty   Yes – some difficulty   Yes – a lot of difficulty   Cannot do at all | |
| Do you have difficulty hearing, even if using a hearing aid? | |  No - no difficulty   Yes – some difficulty   Yes – a lot of difficulty   Cannot do at all | | |  No - no difficulty   Yes – some difficulty   Yes – a lot of difficulty   Cannot do at all | |
| Do you have difficulty walking or climbing steps? | |  No - no difficulty   Yes – some difficulty   Yes – a lot of difficulty   Cannot do at all | | |  No - no difficulty   Yes – some difficulty   Yes – a lot of difficulty   Cannot do at all | |
| Do you have difficulty remembering or concentrating? | |  No - no difficulty   Yes – some difficulty   Yes – a lot of difficulty   Cannot do at all | | |  No - no difficulty   Yes – some difficulty   Yes – a lot of difficulty   Cannot do at all | |
| Do you have difficulty (with self-care such as) washing all over or dressing? | |  No - no difficulty   Yes – some difficulty   Yes – a lot of difficulty   Cannot do at all | | |  No - no difficulty   Yes – some difficulty   Yes – a lot of difficulty   Cannot do at all | |
| Using your usual (customary) language, do you have difficulty communicating,  for example understanding or being understood? | |  No - no difficulty   Yes – some difficulty   Yes – a lot of difficulty   Cannot do at all | | |  No - no difficulty   Yes – some difficulty   Yes – a lot of difficulty   Cannot do at all | |
| **8. Have you made lifestyle changes since your COVID-19 infection? (mark the correct answer with a tick in the box)** | | | | | | |
|  | I do this more often | | I do this less often | No difference | | I did not do this before Covid-19 |
| **Smoking** |  | |  |  | |  |
| **Drinking alcohol** |  | |  |  | |  |
| **Eating healthy food** |  | |  |  | |  |
| **Physical activity**  (including walking & cycling) |  | |  |  | |  |
| **Functioning** \| \| \| \|; **Ability to self-care:** ☐ Same as before COVID-19 ☐ Worse ☐ Better ☐ Unknown \| \| \| \|; **This questionnaire asks about difficulties due to health conditions. Think back over the past 7 days and answer these questions, thinking about how much difficulty you had doing the following activities:** \| **0 No Difficulty** **1 Mild Difficulty** **2 Moderate Difficulty** **3 Severe Difficulty** **4 Extreme Difficulty or Cannot Do** \| **Compared to before COVID-19, are you better/worse/same?** \| \|; \| \| **Better** \| **Worse** \| **Same**; Standing for long periods such as 30 minutes? \| \| \| \|; Taking care of your household responsibilities? \| \| \| \|; Learning a new task, for example, learning how to get to a new place? \| \| \| \|; How much of a problem did you have in joining in community activities (for example, festivities, religious or other activities) in the same way as anyone else can? \| \| \| \|; How much have you been emotionally affected by your health problems? \| \| \| \|; Concentrating on doing something for ten minutes? \| \| \| \|; Walking a long distance such as a kilometre [“or equivalent \| \| \| \|; Washing your whole body? \| \| \| \|; Getting dressed? \| \| \| \|; Dealing with people you do not know? \| \| \| \|; Maintaining a friendship? \| \| \| \|; Your day-to-day work/school? \| \| \| \|; TOTAL \| \| \| \| | | | | | | |

**9. A few questions about your occupation/working status**

**Before you got COVID-19 what was your occupation/working status (paid or unpaid)?**

 Working Full-time  Working Part-time  Full time carer (children or other)  Unemployed

 Unable to work due to chronic illness  Student  Retired  Medically retired  Prefer not to say

**What is your main occupation/working status today?**

 Same as before	 Different from before	 Prefer not to say

**If different, please describe your occupation/working status today (tick all that apply to you)?**

 Working full-time  Working part-time  Not working due to COVID-19 restrictions

 Sick leave  Full time carer (children or others)  Unemployed

 Unable to work due to chronic illness Student  Retired

 Early retirement due to illness  Earning more  Earning less  Prefer not to say

**If different, why did you occupation/working status change?**

 Poor health	 New caring responsibility	 Made redundant

 Working hours reduced by employer	 Sick leave	 Other (specify):

Prefer not to say

**10. A few questions about yourself**

**Sex at Birth:**  Male  Female  Non-binary  Prefer not to say

*~~Ethnicity (tick all that apply):~~* ~~ White  Arab  Black  East Asian  South Asian~~

~~ West Asian  Latin American~~

~~Other (specify):	 Prefer not to say~~ **What is your estimated height:**	. (metres/feet/inches – circle unit used)  Not sure

**What is your current estimated weight:** . (kg/~~Ibs – circle unit used~~)  Not sure

**What was your estimated weight before your Covid19 illness?**	.

(kg/~~Ibs – circle unit used~~)  Not sure

**How many other members regularly live in your household, including yourself:** [_Number_] **What is your highest completed educational level:**

 Primary education(3 to 7 years of school)  Secondary education(8 to 10 years of school)

 Upper Secondary education/High School (11 to 13 years of school)  Vocational / practical school

 Higher College/University  Bachelor degree  Masters degree  PhD

 Other (specify):	 Not completed formal education or training  Prefer not to say

**~~Number of years in formal education:~~**

**11. Please let us know if you feel COVID-19 has affected your health or wellbeing in a way not described above?**

**12. End of survey**

**Thank you for your time!**

### **Table S1**. Categorisation of symptoms at follow-up.

| Symptom category | Long-standing symptoms included |
| --- | --- |
| Respiratory | persistent cough OR shortness of breath/breathlessness OR pain on breathing |
| Gastrointestinal | stomach pain OR nausea OR vomiting OR constipation OR diarrhea |
| Dermatological | lumps or rashes (purple/pink) on toes OR skin rash OR hair loss |
| Chronic fatigue | persistent fatigue OR muscle weakness OR problems sleeping |
| Neurological | tingling feeling/“pins and needles“ OR fainting/ blackouts OR seizures/fits OR tremor/shakiness OR double vision OR problems speaking or communicating OR problems with balance OR confusion/lack of concentration OR dizziness/light headedness OR forgetfulness |
| Mood & Behaviour | anxiety OR loss of interest or pleasure OR behavioural changes OR depressed mood |
| Sensory | loss of smell OR loss of taste |

### **Table S2**. Symptoms reported at the time of the follow-up interview and symptom duration.

| Symptom | Total number of patients with the symptom | Symptom duration | | | | | |
| --- | --- | --- | --- | --- | --- | --- | --- |
|  |  | 1-2 weeks | > 2-4 weeks | >1-2 months | >2-3 months | >3-6 months | From the moment of discharge |
| Persistent dry cough | 177/2623 (6.7%) | 18/2619 (0.69 %) | 9/2619 (0.34%) | 9/2619 (0.34%) | 9/2619 (0.34%) | 17/2619 (0.64%) | 26/2619 (4.24%) |
| Shortness of breath/  breathlessness | 457/2620 (17%) | 12/2614 (0.46%) | 6/2614 (0.23%) | 11/2614 (0.42%) | 9/2614 (0.34%) | 35/2614 (1.34%) | 378/2614 (14.46%) |
| Pain on breathing | 63/2616 (2.4%) | 1/2616 (0.04%) | 3/2616 (0.11%) | 6/2616 (0.22%) | 3/2616 (0.11%) | 6/2616 (0.22%) | 44/2616 (1.68%) |
| Chest pains | 115/2613 (4.4%) | 5/2612 (0.19%) | 9/2612 (0.34%) | 11/2612 (0.42%) | 5/2612 (0.19%) | 9/2612 (0.34%) | 75/2612 (2.9%) |
| Problems with balance | 116/2610 (4.4%) | 5/2606 (0.19%) | 2/2606 (0.08%) | 5/2606 (0.19%) | 7/2606 (0.27%) | 23/2606 (0.88%) | 70/2606 (2.69%) |
| Weakness in arms or legs/ muscle weakness | 269/2598 (10.3%) | 6/2592 (0.23%) | 2/2592 (0.08%) | 7/2592 (0.27%) | 7/2592 (0.27%) | 42/2592 (1.62%) | 199/2592 (7.68%) |
| Swollen ankle(s) | 121/2606 (4.6%) | 6/2602 (0.23%) | 5/2602 (0.19%) | 5/2602 (0.19%) | 6/2602 (0.23%) | 21/2602 (0.8%) | 74/2602 (2.84%) |
| Joint pain or swelling | 174/2600 (6.7%) | 6/2591 (0.23%) | 8/2591 (0.3%) | 4/2591 (0.15%) | 12/2591 (0.46%) | 25/2591 (0.96%) | 110/2591 (4.24%) |
| Can’t fully move or control movement | 43/2602 (1.7%) | 2/2599 (0.08%) | 3/2599 (0.12%) | 1/2599 (0.04%) | 4/2599 (0.15%) | 11/2599 (0.42%) | 18/2599 (0.69%) |
| Persistent muscle pain | 129/2600 (4.97%) | 4/2599 (0.15%) | 7/2599 (0.27%) | 5/2599 (0.19%) | 10/2599 (0.38%) | 19/2599 (0.73%) | 83/2599 (3.19%) |
| Loss of smell | 79/2622 3%) | 3/2622 (0.11%) | 2/2622 (0.08%) | 0/2622 (0%) | 3/2622 (0.11%) | 9/2622 (0.34%) | 62/2622 (2.36%) |
| Loss of taste | 43/2622 (1.6%) | 2/2622 (0.08%) | 1/2622 (0.04%) | 1/2622 (0.04%) | 1/2622 (0.04%) | 5/2622 (0.19%) | 33/2622 (1.26%) |
| Headache | 195/2621 (7.4%) | 27/2616 (1.03%) | 4/2616 (0.15%) | 13/2616 (0.5%) | 7/2616 (0.27%) | 18/2616 (0.69%) | 121/2616 (4.63%) |
| Can’t feel one side of the body or face | 34/2597 (1.3%) | 0/2597 (0%) | 2/2597 (0.08%) | 0/2597 (0%) | 3/2597 (0.12%) | 12/2597 (0.46%) | 17/2597 (0.65%) |
| Tingling feeling/“pins and needles“ | 91/2596 (3.5%) | 9/2593 (0.35%) | 4/2593 (0.15%) | 8/2593 (0.3%) | 8/2593 (0.3%) | 13/2593 (0.5%) | 46/2593 (1.77%) |
| Fainting/ blackouts | 37/2604 (1.4%) | 1/2602 (0.04%) | 1/2602 (0.04%) | 5/2602 (0.19%) | 2/2602 (0.08%) | 7/2602 (0.27%) | 19/2602 (0.73%) |
| Seizures/fits | 60/2601 (2.3%) | 5/2600 (0.19%) | 1/2600 (0.04%) | 4/2600 (0.15%) | 7/2600 (0.27%) | 10/2600 (0.38%) | 32/2600 (1.23%) |
| Tremor/shakiness | 33/2604 (1.2%) | 3/2604 (0.12%) | 2/2604 (0.08%) | 3/2604 (0.12%) | 2/2604 (0.08%) | 3/2604 (0.12%) | 20/2604 (0.77%) |
| Confusion/lack of concentration | 37/2600 (1.42%) | 0/2600 (0%) | 2/2600 (0.08%) | 3/2600 (0.12%) | 2/2600 (0.08%) | 4/2600 (0.15%) | 26/2600 (1%) |
| Problems swallowing or chewing | 21/2601 (0.8%) | 1/2601 (0.04%) | 2/2601 (0.08%) | 2/2601 (0.08%) | 1/2601 (0.04%) | 4/2601 (0.15%) | 11/2601 (0.42%) |
| Double vision | 20/2595 (0.8%) | 0/2594 (0%) | 0/2594 (0%) | 1/2594 (0.04%) | 4/2594 (0.15%) | 4/2594 (0.15%) | 10/2594 (0.39%) |
| Problems speaking or communicating | 20/2601 (0.8%) | 0/2600 (0%) | 1/2600 (0.04%) | 1/2600 (0.04%) | 1/2600 (0.04%) | 4/2600 (0.15%) | 12/2600 (0.46%) |
| Problems sleeping | 264/2589 (10.2%) | 8/2583 (0.3%) | 8/2583 (0.3%) | 12/2583 (0.46%) | 14/2583 (0.54%) | 36/2583 (1.39%) | 180/2583 (6.97%) |
| Dizziness/light headedness | 139/2603 (5.3%) | 14/2601 (0.54%) | 4/2601 (0.15%) | 3/2601 (0.12%) | 7/2601 (0.27%) | 14/2601 (0.54%) | 95/2601 (3.65%) |
| Problems passing urine | 53/2597 (2%) | 5/2597 (0.19%) | 2/2597 (0.08%) | 2/2597 (0.08%) | 3/2597 (0,12%) | 12/2597 (0.46%) | 29/2597 (1.12%) |
| Changes in menstruation | 17/934 (1.8%) | 0/934 (0%) | 1/934 (0.1%) | 2/934 (0.21%) | 2/934 (0.21%) | 4/934 (0.43%) | 8/934 (0.86%) |
| Lumps or rashes (purple/pink) on toes | 5/2599  (0.19%) | 2/2599  (0.08%) | 0/2599  (0%) | 0/2599  (0%) | 1/2599  (0.04%) | 1/2599  (0.04%) | 1/2599  (0.04%) |
| Skin rash | 57/2599  (2.1%) | 6/2597  (0.2%) | 1/2597  (0.04%) | 12/2597  (0.5%) | 4/2597  (0.2%) | 6/2597  (0.2%) | 26/2597  (1.0%) |
| Anxiety | 252/2603  (0.1%) | 3/2600  (0.1%) | 5/2600  (0.2%) | 9/2600  (0.3%) | 12/2600  (0.5%) | 29/2600  (1.1%) | 191/2600  (7.3%) |
| Hair loss | 262/2612  (10.0%) | 1/2580  (0.04%) | 1/2580  (0.04%) | 5/2580  (0.19%) | 14/2580  (0.5%) | 26/2580  (1.0%) | 183/2580  (7.1%) |
| Weight loss | 123/2607  (4.7%) | 4/2604  (0.2%) | 2/2604  (0.08%) | 5/2604  (0.19%) | 8/2604  (0.3%) | 11/2604  (0.4%) | 89/2604  (3.4%) |
| Loss of appetite | 54/2613  (2.1%) | 9/2613  (0.3%) | 5/2613  (0.2%) | 6/2613  (0.2%) | 4/2613  (0.2%) | 8/2613  (0.3%) | 22/2613  (0.8%) |
| Stomach /abdominal pain | 101/2611  (3.9%) | 9/2609  (0.3%) | 2/2609  (0.08%) | 9/2609  (0.3%) | 7/2609  (0.3%) | 10/2609  (0.4%) | 62/2609  (2.4%) |
| Feeling sick | 42/2607  (1.6%) | 6/2605  (0.2%) | 4/2605  (0.2%) | 5/2605  (0.2%) | 3/2605  (0.1%) | 6/2605  (0.2%) | 16/2605  (0.6%) |
| Vomiting | 7/2609  (0.3%) | 1/2608  (0.04%) | 1/2608  (0.04%) | 1/2608  (0.04%) | 1/2608  (0.04%) | 1/2608  (0.04%) | 1/2608  (0.04%) |
| Constipation | 57/2598  (2.2%) | 7/2596  (0.3%) | 7/2596  (0.3%) | 5/2596  (0.2%) | 4/2596  (0.2%) | 5/2596  (0.2%) | 27/2596  (1.0%) |
| Diarrhoea | 66/2601  (2.5%) | 8/2600  (0.3%) | 2/2600  (0.08%) | 4/2600  (0.2%) | 3/2600  (0.1%) | 10/2600  (0.4%) | 38/2600  (1.5%) |
| Palpitations (heart racing) | 186/2607  (7.1%) | 11/2604  (0.4%) | 5/2604  (0.2%) | 9/2604  (0.3%) | 12/2604  (0.5%) | 19/2604  (0.7%) | 127/2604  (4.9%) |
| Bleeding | 18/2597  (0.7%) | 3/2597  (0.1%) | 4/2597  (0.2%) | 1/2597  (0.04%) | 2/2597  (0.08%) | 3/2597  (0.1%) | 5/2597  (0.2%) |
| Persistent fatigue | 656/2609  (25.1%) | 10/2599  (0.4%) | 5/2599  (0.2%) | 15/2599  (0.6%) | 12/2599  (0.5%) | 52/2599  (2.0%) | 551/2599  (21.2%) |
| Stiffness of muscles, rigidity | 58/2598  (2.2%) | 2/2596  (0.08%) | 1/2596  (0.04%) | 6/2596  (0.2%) | 2/2596  (0.08%) | 10/2596  (0.4%) | 35/2596  (1.3%) |
| Slowness of movement (bradykinesia) | 65/2599  (2.5%) | 2/2597  (0.08%) | 1/2597  (0.04%) | 3/2597  (0.1%) | 6/2597  (0.2%) | 12/2597  (0.5%) | 39/2597  (1.5%) |
| Problems seeing | 280/2603  (10.8%) | 3/2598  (0.1%) | 4/2598  (0.2%) | 11/2598  (0.4%) | 16/2598  (0.6%) | 43/2598  (1.7%) | 198/2598  (7.6%) |
| Problems hearing | 133/2602  (5.1%) | 4/2597  (0.2%) | 4/2597  (0.2%) | 5/2597  (0.2%) | 6/2597  (0.2%) | 26/2597  (1.0%) | 83/2597  (3.2%) |
| Forgetfulness | 296/2600  (11.4%) | 3/2597  (0.1%) | 4/2597  (0.2%) | 8/2597  (0.3%) | 8/2597  (0.3%) | 33/2597  (1.3%) | 237/2597  (9.1%) |
| Worsening weakness and/or respiratory function in patients with pre-existing neurological or neuromuscular condition | 29/2588  (1.1%) | 1/2587  (0.04%) | 2/2587  (0.08%) | 0/2587  (0%) | 2/2587  (0.08%) | 5/2587  (0.2%) | 18/2587  (0.7%) |
| Loss of interest or pleasure | 152/2590  (5.9%) | 2/2586  (0.08%) | 3/2586  (0.1%) | 5/2586  (0.2%) | 7/2586  (0.3%) | 13/2586  (0.5%) | 118/2586  (4.6%) |
| Behavioural changes | 103/2592  (3.97%) | 0/2589  (0%) | 2/2589  (0.08%) | 4/2589  (0.2%) | 5/2589  (0.2%) | 14/2589  (0.5%) | 75/2589  (2.9%) |
| Depressed mood | 169/2595  (6.5%) | 3/2588  (0.1%) | 4/2588  (0.2%) | 6/2588  (0.2%) | 6/2588  (0.2%) | 17/2588  (0.7%) | 125/2588  (4.8%) |
| Hallucinations (seeing or hearing things others don’t see or hear) | 12/2593  (0.5%) | 1/2593  (0.04%) | 0/2593  (0%) | 0/2593  (0%) | 0/2593  (0%) | 1/2593  (0.04%) | 10/2593  (0.4%) |
| Hyperhidrosis | 161/2611  (6.2%) | 2/2591  (0.08%) | 5/2591  (0.2%) | 4/2591  (0.2%) | 4/2591  (0.2%) | 10/2591  (0.4%) | 116/2591  (4.5%) |
| Myoclonus | 14/1319  (1.1%) | 0/1319  (0%) | 0/1319  (0%) | 2/1319  (0.2%) | 5/1319  (0.4%) | 2/1319  (0.2%) | 5/1319  (0.4%) |

### **Table S3**. Perceived breathlessness post- and pre-COVID-19 assessed using MRC Dyspnoea Scale at the time of the follow-up.

|  | Grade | Degree of breathlessness related to activity | Total |
| --- | --- | --- | --- |
| Within the last 24 hours | 1 | 1, Not troubled by breathlessness except on strenuous exercise | 1635 (61.7%) |
|  | 2 | 2, Short of breath when hurrying or when walking up a slight hill | 617 (23.3%) |
|  | 3 | 3, I walk slower than most people of my age because of breathlessness, or have to stop for breath when walking at own pace | 194 (7.3%) |
|  | 4 | 4, I stop for breath after walking 90-100 meters, or after a few minutes on level ground | 93 (3.5%) |
|  | 5 | 5, Too breathless to leave the house, or breathless when dressing/undressing | 31 (1.2%) |
|  | NA | Unknown | 79 (3%) |
| Before  COVID-19 illness | 1 | 1, Not troubled by breathlessness except on strenuous exercise | 1970 (74.3%) |
|  | 2 | 2, Short of breath when hurrying or when walking up a slight hill | 461 (17.4%) |
|  | 3 | 3, I walk slower than most people of my age because of breathlessness, or have to stop for breath when walking at own pace | 87 (3.3%) |
|  | 4 | 4, I stop for breath after walking 90-100 meters, or after a few minutes on level ground | 37 (1.4%) |
|  | 5 | 5, Too breathless to leave the house, or breathless when dressing/undressing | 15 (0.6%) |
|  | NA | Unknown | 79 (3%) |

### **Table S4**. EuroQol visual analog scale asking participants to score health state from 0 (worst imaginable health) to 100 (best imaginable health) before COVID-19 and at the time of the follow-up, stratified by symptom categories.

| Category  of long-standing symptoms (n) | Before Covid, median (IQR) | At the time of the follow-up, median (IQR) | p-value  Pre-Covid  vs  No Symptoms | p-value  At the time of the follow-up, vs  No symptoms | p-value  At the time of the follow-up vs  Pre-Covid |
| --- | --- | --- | --- | --- | --- |
| No symptoms (1115) | 90 (76.5-99.5) | 85 (75-95) |  |  | 0.11 |
| Respiratory (n=451) | 80 (70-90) | 70 (50-80) | **<0.001** | **<0.001** | **<0.001** |
| Gastrointestinal (n=110) | 80 (70-88) | 65 (50-75) | **<0.001** | **<0.001** | **<0.001** |
| Dermatological (n=206) | 83 (75-95) | 70 (60-87) | 0.1 | **<0.001** | **<0.001** |
| Chronic fatigue (n=658) | 80 (70-90) | 70 (50-80) | **<0.001** | **<0.001** | **<0.001** |
| Neurological (n=375) | 80 (70-90) | 69.5 (50-80) | **<0.001** | **<0.001** | **<0.001** |
| Mood & Behaviour (n=284) | 80 (70-90) | 67 (50-75) | **<0.001** | **<0.001** | **<0.001** |
| Sensory (n=70) | 86 (80-91.3) | 75 (69-85) | 0.65 | **<0.001** | **<0.001** |

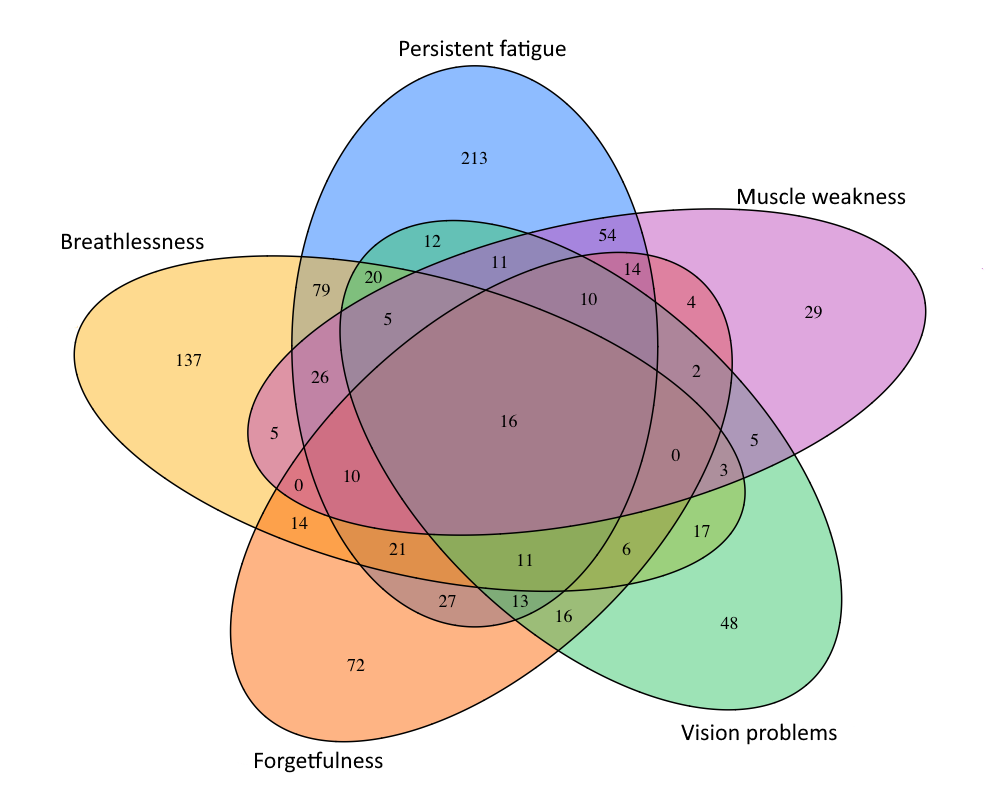

**A**

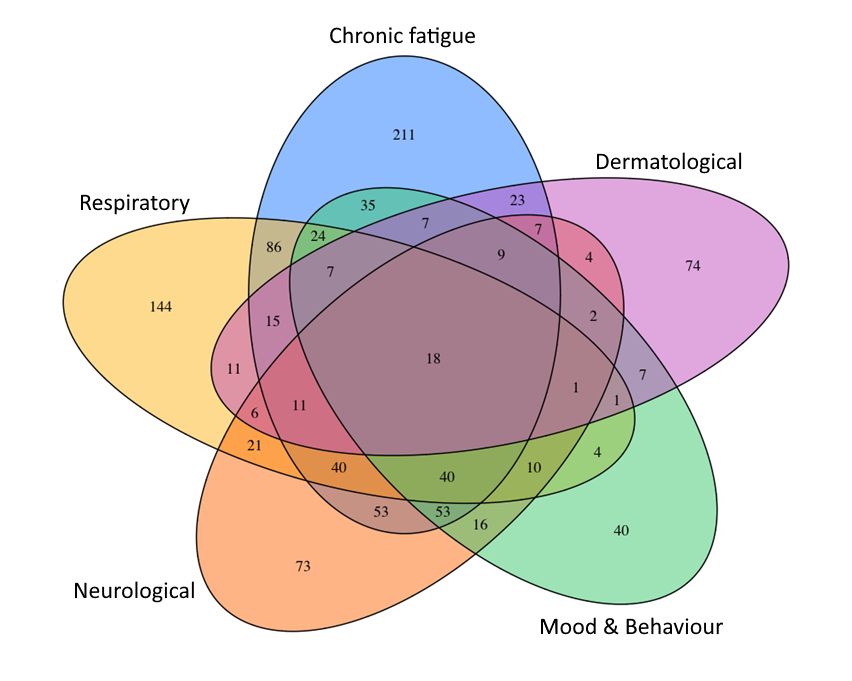

**B**

### Figure S1. Venn plot presenting coexistence of (a) five most common long-standing symptoms and (b) five most common categories of long-standing symptoms at the time of the follow-up interview.

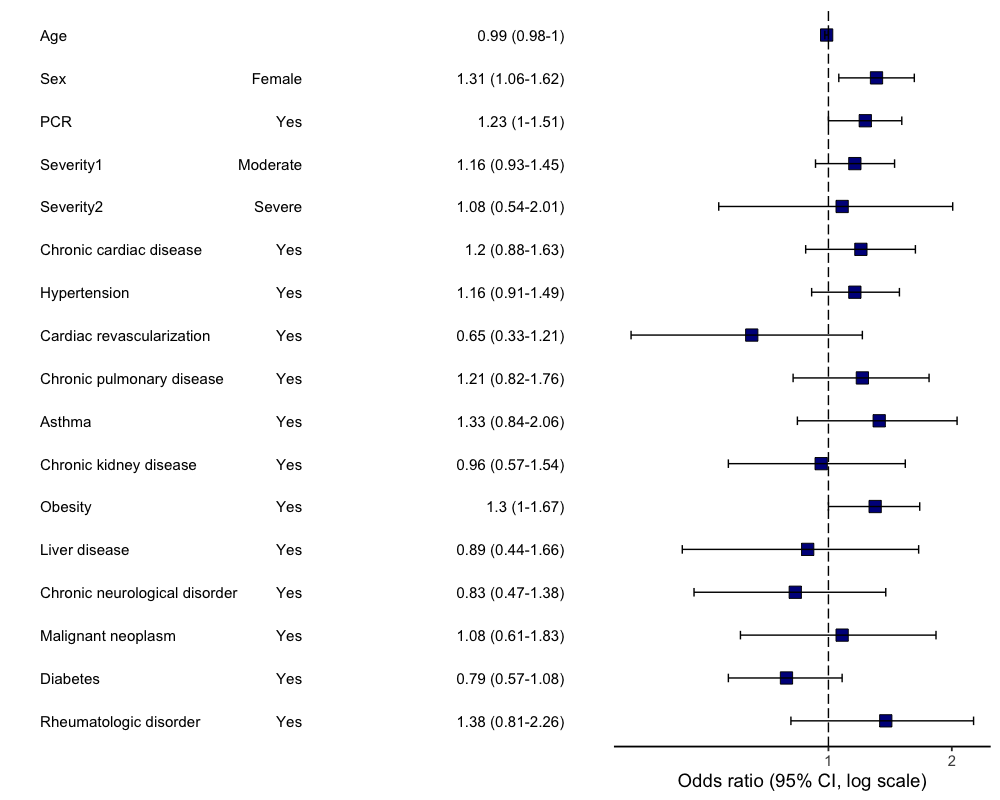

**B**

**A**

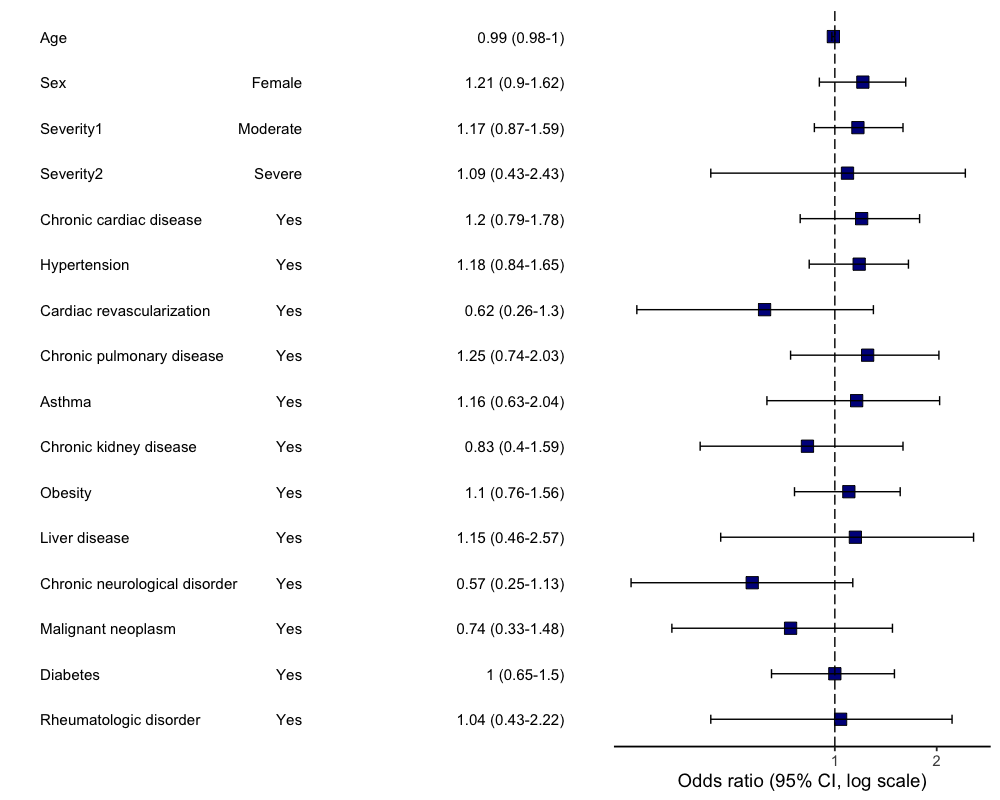

Figure S2. Multivariable logistic regression model. Odds ratios and 95% CIs for “Respiratory” category of long-standing symptoms at the time of follow-up. Abbreviation: CI, confidence interval. (a) primary analysis (age, sex, comorbidities, severity and RT-PCR were included as potential risk factors); (b) sensitivity analysis (performed in a subgroup of RT-PCR positive patients only).

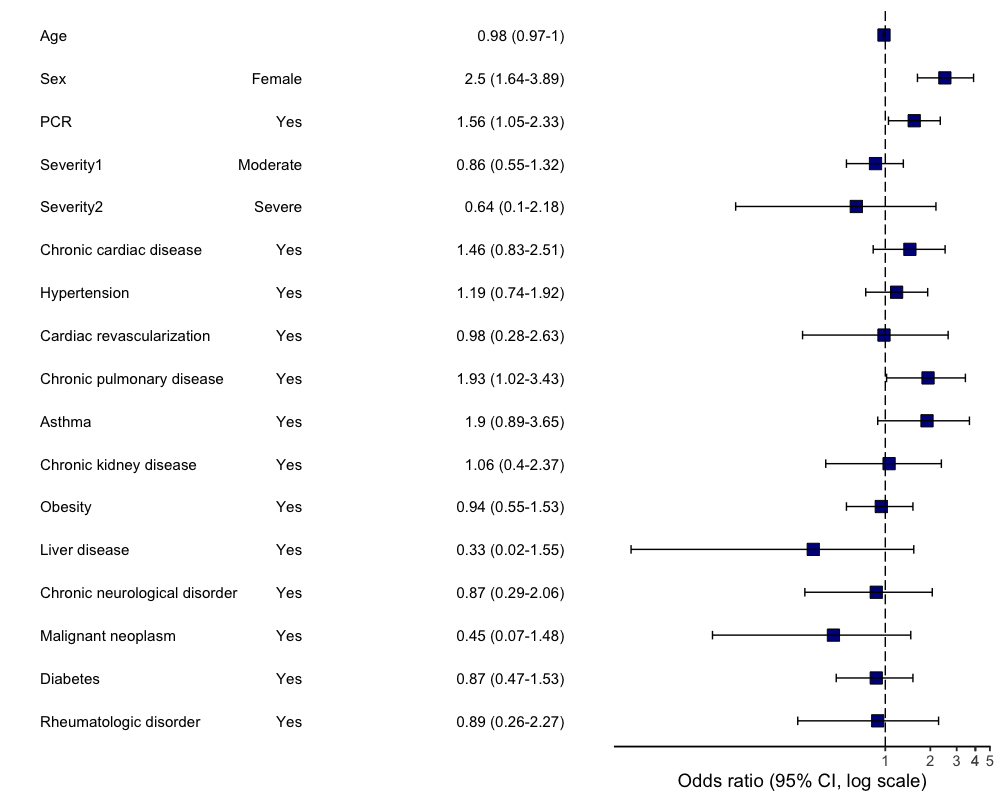

**B**

**A**

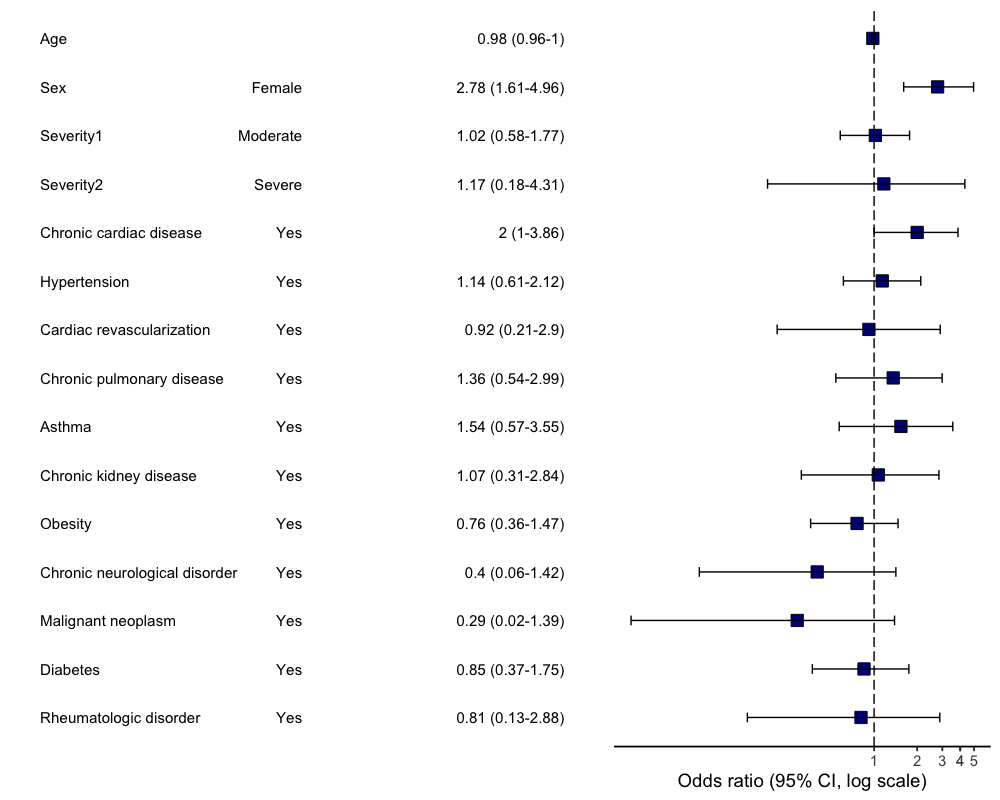

Figure S3. Multivariable logistic regression model. Odds ratios and 95% CIs for “Gastrointestinal” category of long-standing symptoms at the time of follow-up. Abbreviation: CI, confidence interval. (a) primary analysis (age, sex, comorbidities, severity and RT-PCR were included as potential risk factors); (b) sensitivity analysis (performed in a subgroup of RT-PCR positive patients only).

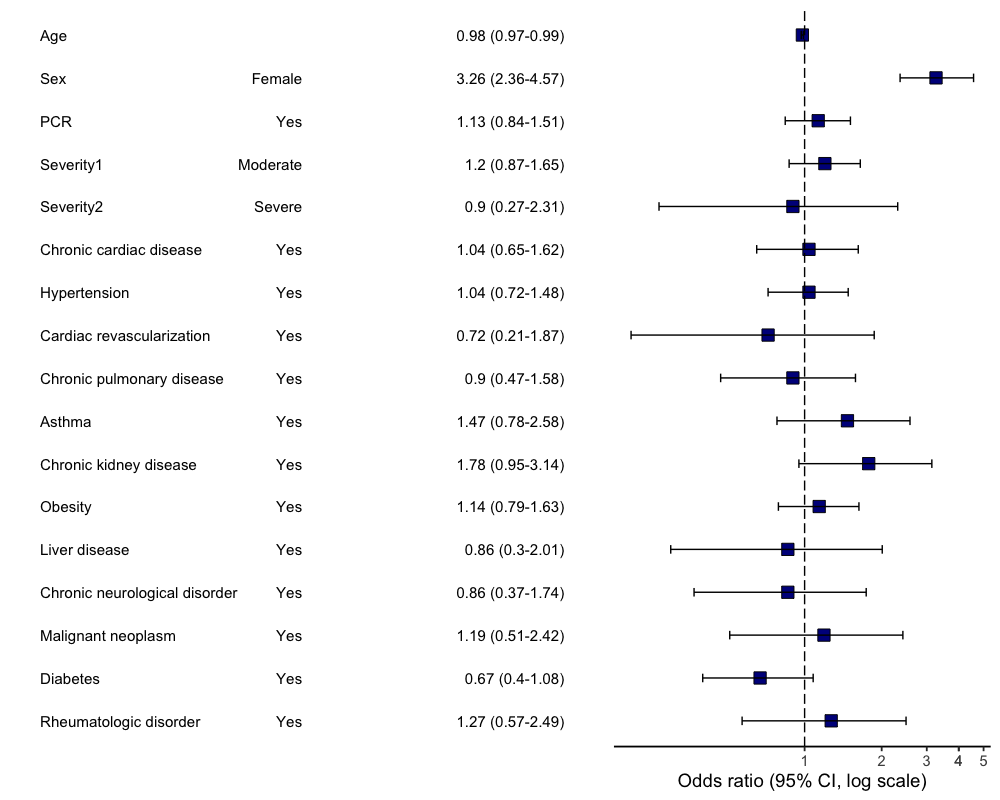

**B**

**A**

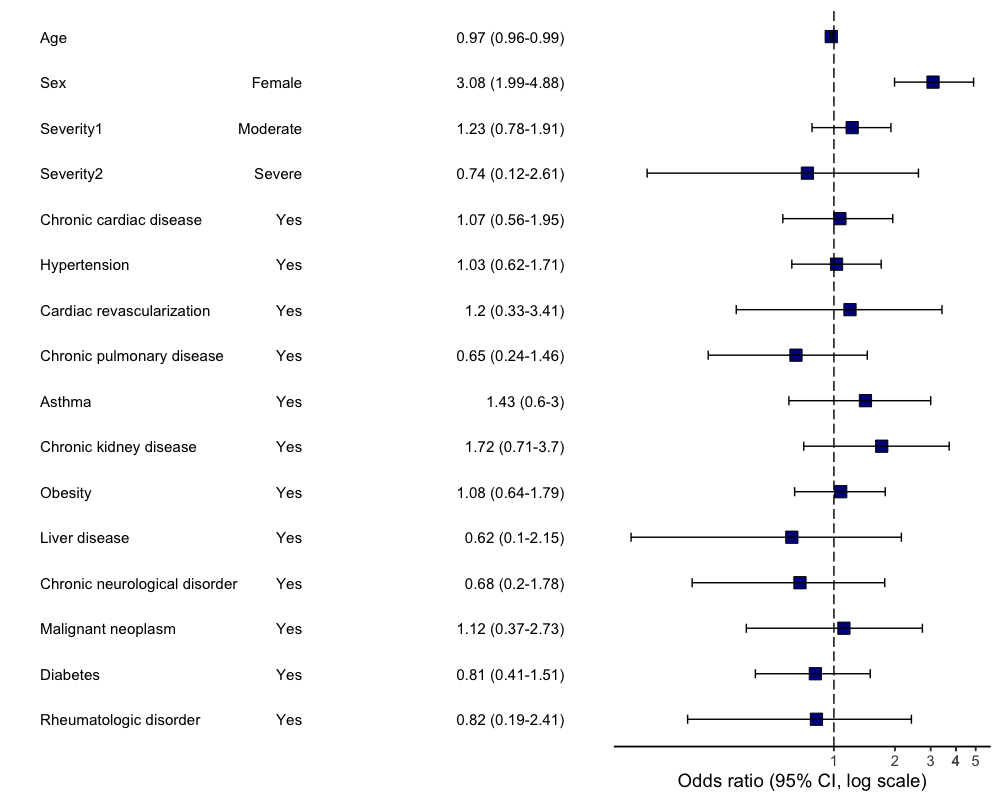

Figure S4. Multivariable logistic regression model. Odds ratios and 95% CIs for “Dermatological” category of long-standing symptoms at the time of follow-up. Abbreviation: CI, confidence interval. (a) primary analysis (age, sex, comorbidities, severity and RT-PCR were included as potential risk factors); (b) sensitivity analysis (performed in a subgroup of RT-PCR positive patients only).

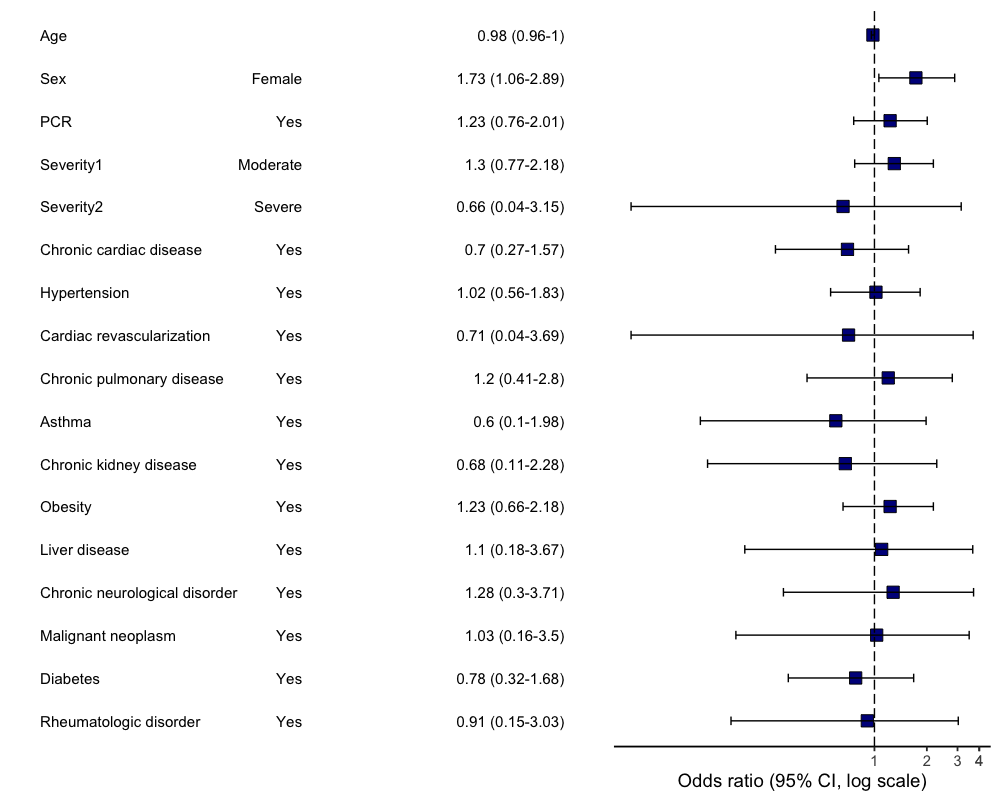

**B**

**A**

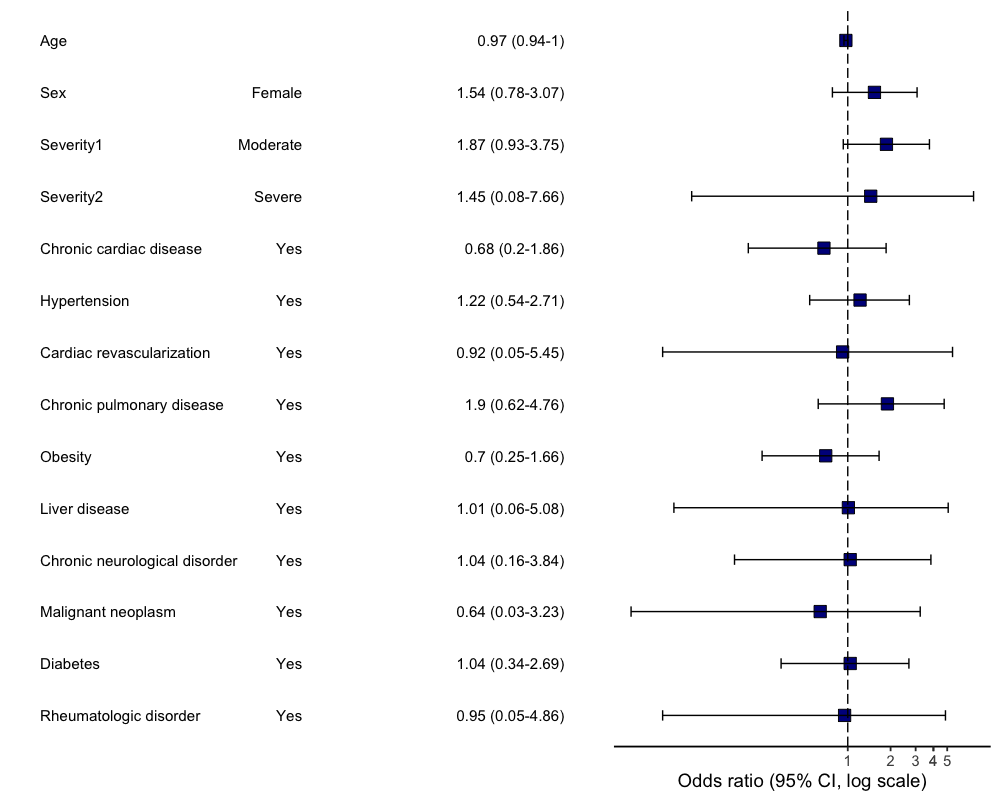

Figure S5. Multivariable logistic regression model. Odds ratios and 95% CIs for “Sensory” category of long-standing symptoms at the time of follow-up. Abbreviation: CI, confidence interval. (a) primary analysis (age, sex, comorbidities, severity and RT-PCR were included as potential risk factors); (b) sensitivity analysis (performed in a subgroup of RT-PCR positive patients only).

1. the CRF parts taken from **The WHO Global Neuro COVID-19 Clinical Case Record Form** presented in the grey shading

   information added to the ISARIC CRF by the StopCOVID Team is presented in red font

   Questions not asked during telephone interview are highlighted by strikethrough
